# A multimorphic variant in ThPOK causes a novel human disease characterized by T cell abnormalities, immunodysregulation, allergy, and fibrosis

**DOI:** 10.1101/2024.06.26.24309360

**Authors:** Maryam Vaseghi-Shanjani, Mehul Sharma, Pariya Yousefi, Simran Samra, Kaitlin U. Laverty, Arttu Jolma, Rozita Razavi, Ally H. W. Yang, Mihai Albu, Liam Golding, Anna F. Lee, Ryan Tan, Phillip A. Richmond, Marita Bosticardo, Jonathan H. Rayment, Connie L. Yang, Kyla J. Hildebrand, Rae Brager, Michelle K. Demos, Yu Lung Lau, Luigi D. Notarangelo, Timothy R. Hughes, Catherine M. Biggs, Stuart E. Turvey

## Abstract

ThPOK is best known as a regulator of CD4+ T cell lineage commitment, although it was initially cloned as a suppressor of collagen expression in the skin. The role of ThPOK has not been formally established in humans since individuals with damaging variants in ThPOK have not yet been identified. Here, we report the first case of a human with a damaging heterozygous *de novo* variant in ThPOK causing a syndrome encompassing CD4+ T cell deficiency, allergy, fibroinflammatory interstitial lung disease, developmental delay, and growth failure. The patient’s variant, ThPOK^K360N^, exhibited abnormal multimorphic activity, including interfering with ThPOK^WT^ in regulating gene regulation (antimorph). Protein-DNA interaction assays showed inability of ThPOK^K360N^ to bind to wild-type consensus sequences (amorph) and revealed a novel DNA-binding specificity (neomorph). Single-cell RNA sequencing of peripheral blood revealed potential developmental defects in maturation and activation of CD4+ and CD8+ T cells (hypomorph). To establish causality, we recapitulated the observed cellular defects in lentivirally transduced healthy control T cells and pulmonary fibroblasts. Transcriptomic analysis showed that T cells transduced with ThPOK^K360N^ lacked the upregulation of activation, proliferation, and functional pathways observed in ThPOK^WT^-transduced cells. When overexpressed in healthy control fibroblasts, ThPOK^K360N^ significantly increased the expression of pro-fibrotic genes implicated in pulmonary fibrosis, indicating a defect in regulating collagen expression. This novel human disease caused by a multimorphic variant in ThPOK confirms its role in CD4+ T cell development in an intact human context, while also revealing an unanticipated role for ThPOK in T cell function and the regulation of fibrotic pathways in fibroblasts.

**One sentence summary:** Discovery of a human with pathogenic multimorphic genetic change in ThPOK establishes ThPOK as essential for human T cell development and reveals its novel role as a regulator of fibrosis.

## INTRODUCTION

T-helper-inducing POZ/Kruppel-like factor (ThPOK), encoded by *ZBTB7B*, is a transcription factor that belongs to the zinc finger and BTB/POZ domain-containing (ZBTB) family of proteins (*1*). ThPOK acts both as an activator and a suppressor of transcription through direct binding to promoter regions of target genes and by interaction with various co-regulators (*2*). In recent years, ThPOK has garnered significant attention in immunological research, primarily through insights gained from murine models. The most well-studied function of ThPOK is its role in directing the differentiation of T cell lineages, particularly in promoting the development of CD4+ T cells and inhibiting the development of CD8+ T cells in the thymus (*3*). This discovery was initially made based on a spontaneous mutant mouse strain with biallelic missense variants in the second zinc finger domain of ThPOK. These so-called ‘helper-deficient’ (HD) mice show a selective absence of mature CD4+ T cells and increased frequency of CD8+ T cells. While the primary focus on ThPOK has been its immune functions, its role extends to other biological processes, although these are less well-understood. For example, ThPOK is highly expressed in the skin and downregulates type I collagen gene expression in murine skin fibroblasts, binding specifically to the regulatory regions of type I collagen genes (*Col1a1* and *Col1a2*) (*4*).

Despite advances in our understanding of the role of ThPOK in immunity, the direct implications of *ZBTB7B* variants in human health have remained largely unexplored. To date, disabling genetic changes in ThPOK have not been implicated as a cause of human monogenic disease. Here, we describe a patient with a *de novo* heterozygous pathogenic variant in *ZBTB7B* causing a complex syndromic phenotype including CD4+ T lymphopenia, CD8+ T lymphocytosis, early-onset allergic disease, severe fibroinflammatory interstitial lung disease, and failure to thrive. Our detailed mechanistic investigations reveal that this variant is multimorphic, exhibiting antimorphic dominant-negative (DN), amorphic and hypomorphic loss-of-function (LOF), and neomorphic gain-of-function (GOF) effects. Our findings, in the context of the whole human organism, both reinforce the well-established role of ThPOK in T cell development, as demonstrated by the patient’s abnormalities in T cell development and function, but also emphasize the unexpected role of ThPOK in pulmonary fibrosis, as demonstrated by the increase in pro-fibrotic gene signature in fibroblasts expressing the variant. These findings not only elucidate and challenge the current understanding of ThPOK’s role in the immune system, but also open new avenues for exploring its broader implications in other tissues and body systems in humans.

## RESULTS

### A novel *de novo* heterozygous *ZBTB7B* variant in a child with immunodeficiency, allergic disease, and interstitial lung disease

The patient is young male child born to healthy non-consanguineous parents. His complex clinical course was notable for global developmental delay, failure to thrive (Figure S1), bilateral sensorineural hearing loss (Figure S2), non-healing corneal ulceration, increased echogenicity of multiple tissues (i.e. pancreas, liver and kidneys), growth hormone deficiency, allergic disease, combined immunodeficiency, and severe interstitial lung disease requiring supplemental oxygen (Figure 1A). He had persistent chest imaging abnormalities (Figure 1B-C) and lung biopsy demonstrated chronic inflammation and fibrosis of the lung, characterized by lipoproteinosis and notable subpleural cystic remodeling and honeycomb changes (Figure 1D-I).

**Figure 1.**
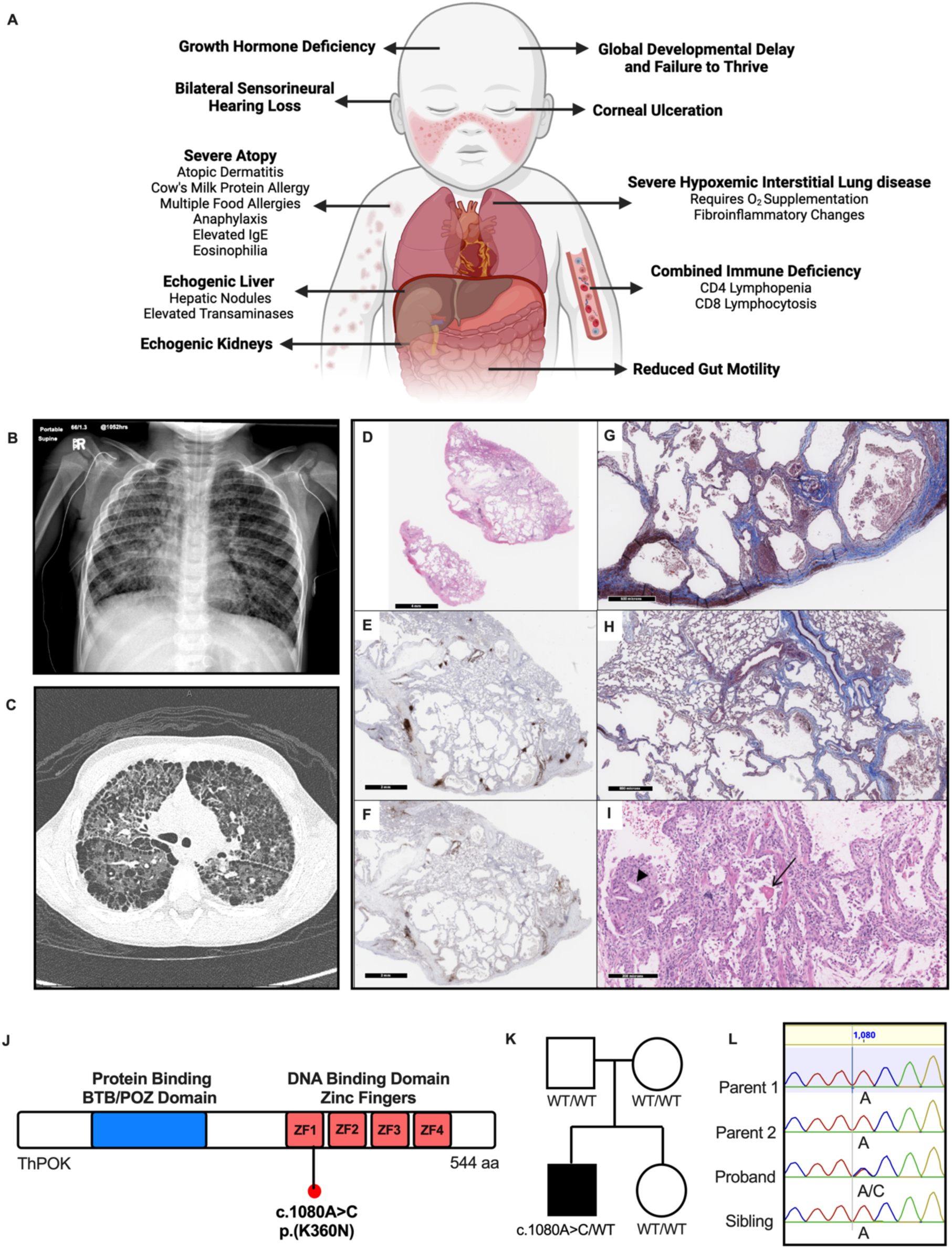
Trio genome sequencing revealed a *de novo* heterozygous missense variant in *ZBTB7B* in a pediatric patient with a multi-system disorder. **(A)** A summary of patient’s clinical features. **(B)** Chest x-ray showing hyperinflated lungs, coarse reticular shadowing, and interstitial septal thickening. **(C)** Chest CT (coronal view) showing ground glass opacity and subpleural honeycombing changes. **(D)** Low-power view lung wedge biopsy (H&E stain). **(E)** CD20 immunohistochemistry highlights (in brown) the B cell population of the lymphoid aggregates in the interlobular septa. **(F)** CD3 immunohistochemistry highlights (in brown) the T cell population of the lymphoid aggregates in the interlobular and alveolar septa. **(G)** Subpleural honeycomb change is evident on trichrome stain, which highlights the interstitial fibrosis in blue. **(H)** The lung architectural abnormalities are patchy (non-uniform), as demonstrated by relatively normal parenchyma in the top half of this image, and interstitial fibrosis and alveolar widening in the lower half (trichrome stain). The patchy changes can also be appreciated in the low-power photomicrographs. **(I)** Some alveoli contain aggregates of alveolar macrophages surrounding cholesterol clefts (arrowhead), as well as amorphous eosinophilic material (arrow) (H&E stain). All images have scale bars embedded in the respective panels. **(J)** Illustration of the ThPOK protein, with four C-terminal zinc fingers responsible for DNA-binding to the consensus sequence and the N-terminal protein-binding BTB/POZ domain. Location of the variant (NM_001256455.2:c.1080A>C, p.Lys325Asn) is shown (red dot). ZF, zinc finger. **(K)** Family pedigree of the patient. Filled symbols = affected individuals; unfilled symbols = unaffected individuals. WT, wild-type. **(L)** Sanger sequencing results of all family members indicating heterozygous substitution at position 1080 found only in the proband.

Although T cell receptor excision circle (TREC) counts were normal at birth, CD4+ lymphopenia was identified in infancy. Detailed immunophenotyping showed CD4+ lymphopenia and reduced naïve CD4+ T cell percentage, with the opposite finding in CD8+ T cells of lymphocytosis and elevated naïve percentage, and B cell lymphocytosis (Table S1).

Due to this unusual constellation of issues, trio whole-genome sequencing was pursued which identified a novel heterozygous *de novo* variant in *ZBTB7B* (NM_001256455.2:c.1080A>C, p.K360N), predicted to be damaging using *in silico* prediction tools (Table S2). The sequencing results were validated by Sanger sequencing in all family members (Figure 1J-L). The variant results in a substitution from a positively charged lysine residue to a polar uncharged asparagine residue at a highly conserved region of the protein (Figure S3), altering the first zinc finger in the DNA-binding domain of ThPOK (Figure 1J).

### ThPOK^K360N^ displays altered neomorphic DNA binding specificity

We assessed the effect of the NM_001256455.2:c.1080A>C (p.K360N) variant on protein expression using HEK293 cells. We selected HEK293 cells as our model system as these cells lack endogenous expression of ThPOK (www.proteinatlas.org/). Expression of ThPOK^K360N^ was assessed both in isolation and in combination with ThPOK^WT^, modelling the patient’s heterozygous state. Expression of ThPOK^K360N^ remained comparable to ThPOK^WT^ both in isolation and in the heterozygous state, suggesting that the presence of the variant does not impact protein expression (Figure 2A, Figure S4).

**Figure 2.**
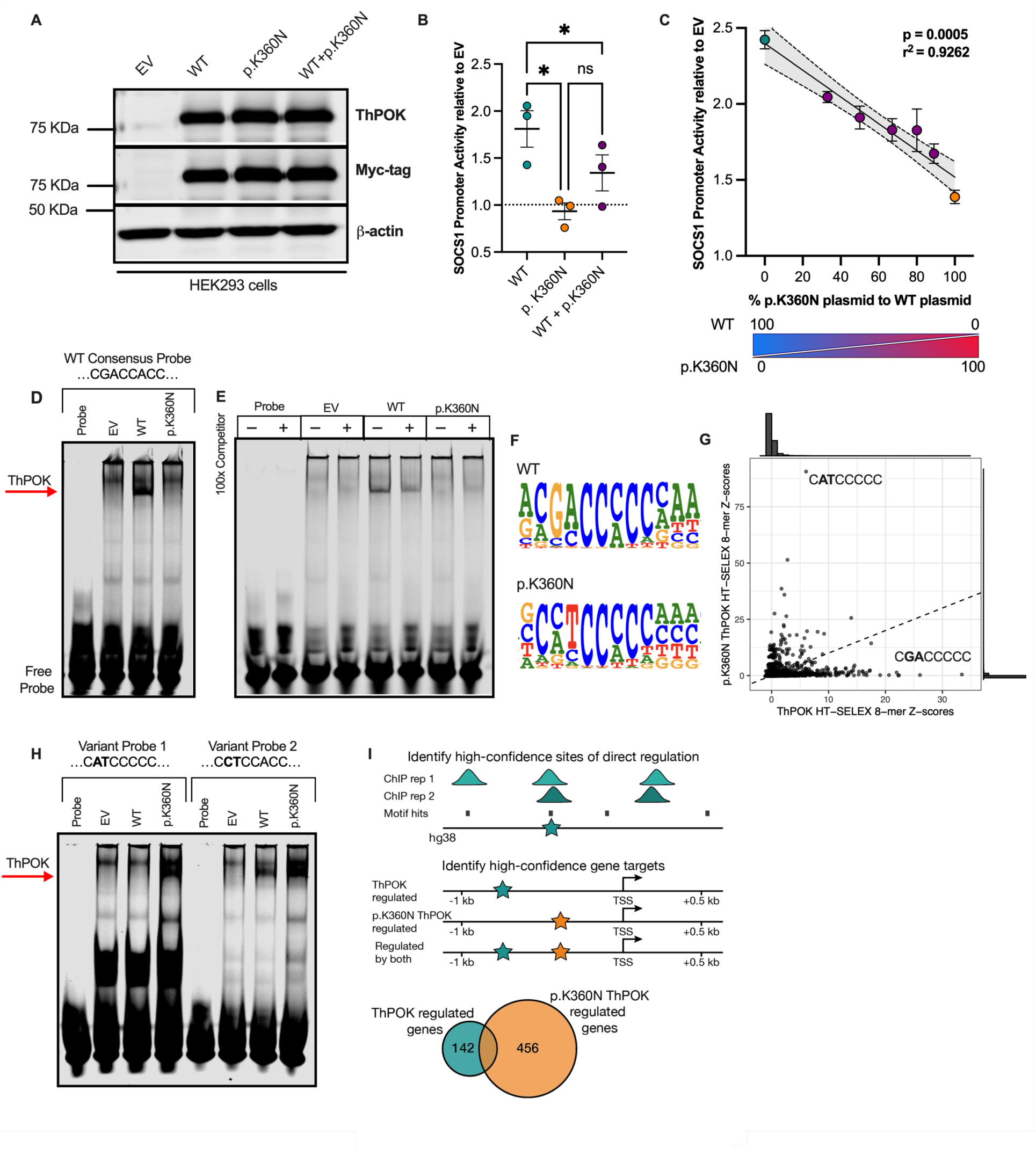
ThPOK^K360N^ showed altered DNA-binding specificity. **(A)** Comparative analysis of ThPOK^K360N^ and ThPOK^WT^ protein expression using immunoblotting in HEK293 cells. Protein was detected using anti-ThPOK and anti-Myc antibodies. β-actin was assessed as loading control. Representative blot from three independent experiments. **(B-C)** Luciferase reporter assay using a region of *SOCS1* promoter in HEK293 cells transfected with various combinations and ratios of vectors indicated below the plot. Firefly luciferase activity was normalized to Renilla luciferase to account for transfection efficiency variations, with values further normalized to the empty vector (EV) condition, offering a relative comparison to the EV baseline. Data are presented as mean ± SEM, derived from three independent biological replicates, each comprising of the average of three technical replicates. For panel B, statistical analysis was conducted using one-way ANOVA for repeated measures, with Sidak’s method applied for multiple comparisons correction. For panel C, dotted lines represent the 95% confidence intervals for the Pearson correlation coefficient. **(D)** EMSA showing *in vitro* DNA binding of ThPOK^WT^ and ThPOK^K360N^. Whole cell lysates of HEK293 cells transfected with an expression vector encoding each protein were incubated with infrared fluorescent dye labeled *SOCS1* probe. Anti-ThPOK antibody generated a supershift of the DNA–protein complexes (red arrow). Representative image from three independent experiments. **(E)** Competitor EMSA assay using an unlabeled (cold) probe identical to the labelled probe sequence. **(F)** Position weight matrices (PMW) for ThPOK^WT^ and ThPOK^K360N^, identified from HT-SELEX data, showing the relative frequency of nucleotide occurrence at each position within the binding site. **(G)** Relative binding affinities of ThPOK^WT^ and ThPOK^K360N^ to all 8-base long DNA sequences (8-mers). Each point on the plot represents a unique 8-mer, with Z-scores calculated from the number of occurrences of the 8-mer in bound DNA sequences in the third HT-SELEX cycle. The sequence of top 8-mer for each protein variant is shown, as well as marginal distributions of 8-mer Z-scores (top and right side of the scatter plot). **(H)** EMSA showing *in vitro* DNA binding of ThPOK^WT^ and ThPOK^K360N^ to variant probes designed using binding sequences revealed by HT-SELEX. **(I)** Schematic depicting identification of ThPOK^WT^ and ThPOK^K360N^ regulated genes, i.e., genes with a high-confidence binding site of the protein variant within its promoter region. For each protein variant, high confidence binding sites were identified as loci with a PWM hit (using HT-SELEX derived motifs) that overlapped a peak from each ChIP-seq replicate. These loci were mapped to promoter regions 1 kb upstream to 0.5 kb downstream of a transcription start site (TSS) to identify genes that are likely regulated by each protein variant. Numbers in the Venn diagram show the total number of genes regulated by ThPOK^WT^ and ThPOK^K360N^, 36 of which are shared between both.

Next, we evaluated the effect of the variant on the function of ThPOK as a transcription factor and its regulatory influence on *SOCS1* expression. Building on Luckey *et al*.’s identification of ThPOK-binding sites within the mouse *Socs1* promoter region (*5*), we developed a luciferase reporter assay utilizing a segment of the human *SOCS1* promoter containing the previously identified DNA binding motif of ThPOK (*6*). We established that ThPOK^K360N^ is impaired in its ability to upregulate *SOCS1* transcriptional activity compared to ThPOK^WT^ (Figure 2B). When co-transfecting cells to mimic the heterozygous state with both wild-type (WT) and p.K360N *ZBTB7B* plasmids, the transcriptional activity of *SOCS1* exhibited an intermediate increase (Figure 2B). We then investigated the potential dominant-negative effect of ThPOK^K360N^ on ThPOK^WT^ by systematically increasing the ratio of the variant plasmid relative to the WT plasmid. Here we observed an inverse relationship between the amount of the variant plasmid and *SOCS1* transcriptional activity (Pearson correlation: r^2^ = 0.9262, p-value = 0.0005) (Figure 2C).

To further assess the DNA-binding ability of ThPOK^K360N^, we employed an electrophoretic mobility shift assay (EMSA) using a tagged 35 nucleotide probe from the *SOCS1* promoter. We observed that ThPOK^WT^ could effectively bind to this probe, while ThPOK^K360N^ could not (Figure S5). To ensure the observed binding was specific, a supershift assay was performed and confirmed that the ThPOK variant exhibited no detectable binding activity, indicating a loss of DNA-binding capability likely attributable to the missense change in the DNA-binding zinc finger (Figure 2D). Additionally, a competition assay with a 100-fold excess of unlabeled competitor probe substantially diminished the supershifted ThPOK^WT^-DNA complex band, further verifying the specificity of the binding (Figure 2E).

We used structural modeling to evaluate the predicted impact of the amino acid change at position 360 on the ability of ThPOK to interact with DNA, using the paralog ZBTB7A (PDB:8E3D), which shares sequence homology in the DNA-binding zinc finger domain with ThPOK (*7*). The structural analysis of ZBTB7A bound to a DNA target revealed that the lysine residue, equivalent to the changed 360 site in ThPOK, forms direct hydrogen bonds with the bases of AG dinucleotide. Consequently, the variant in ThPOK likely disrupts these interactions, altering its DNA binding specificity (Figure S6). To experimentally define and compare the DNA binding profiles of ThPOK^WT^ and ThPOK^K360N^, we utilized high-throughput systematic evolution of ligands by exponential enrichment (HT-SELEX). The HT-SELEX results further corroborated changes in DNA binding specificity of ThPOK^K360N^ compared to ThPOK^WT^ (Figure 2F). Position weight matrices (PWM) revealed a shift in nucleotide preference in positions 3 and 4, specifically changing ‘GA’ nucleotides, preferred by the WT, to ‘CT’ or ‘AT’ nucleotides, preferred by ThPOK^K360N^. Notably, both ‘C’ and ‘A’ were similarly preferred at position 3 for ThPOK^K360N^ (Figure 2F). Additionally, the relative abundance of 8-mer sequences from HT-SELEX experiments revealed that sequences with high affinity for either ThPOK^WT^ or ThPOK^K360N^ generally had a low incidence in the counterpart’s selection. This distinct separation underscores the divergent DNA binding specificities between ThPOK^WT^ and ThPOK^K360N^ (Figure 2G).

Given that the original ’GA’ dinucleotide at positions 3 and 4 in the WT binding sequence is highly conserved and exhibits low flexibility, changes to this sequence are likely to impact binding specificity. To test this using EMSA, we created two variant oligonucleotide probes informed by the highest hit of the 8-mer plot (Variant Probe 1) and the consensus sequence for ThPOK^K360N^ as indicated by PMW obtained from HT-SELEX (Variant Probe 2) that incorporated the dinucleotide modifications ‘AT’ (3’…C**AT**CCCCC…5’) or ‘CT’ (3’…C**CT**CCACC…5’) respectively (see Table S3 for full-length probe sequences). Our results demonstrated that ThPOK^K360N^ exhibited strong binding affinity to Variant Probe 1 and 2 (Figure 2H).

To further assess the DNA binding landscape of the transcription factor, we integrated the HT-SELEX motifs with corresponding chromatin immunoprecipitation followed by sequencing (ChIP-seq) data to identify high-confidence gene targets for both ThPOK^WT^ and ThPOK^K360N^. A gene was designated as a high-confidence target based on the co-occurrence of ChIP-seq peaks in all replicates within the promoter regions (spanning from −1000 to +500 bp relative to the transcription start site) that also harbored corresponding HT-SELEX motif hits (Figure 2I). Our analysis revealed distinct gene targets likely to be regulated by ThPOK^WT^ and ThPOK^K360N^, as illustrated by the Venn diagram (Figure 2I), with limited overlap between the gene sets and associated pathways (data file S1) targeted by each protein, suggesting that ThPOK^WT^ and ThPOK^K360N^ engage in differential regulatory networks. This pattern indicates potentially unique transcriptional roles for each protein, which may underlie differences in cellular responses and phenotypic outcomes.

### Patient T cells have impaired development, memory phenotype skewing, and enhanced Th2 effector function

ThPOK plays a critical role in the development of lymphoid cells, particularly in the lineage commitment of CD4+ T cells (*1–3, 5, 8*) (Figure 3A). To define how ThPOK^K360N^ impacts T cell development, we first performed immunophenotyping of the patient’s lymphoid compartment. Clinical flow cytometry data, collected over multiple time points, consistently revealed T cell abnormalities; notably persistent CD4+ lymphopenia, CD8+ lymphocytosis, and a lower CD4+/CD8+ ratio, all while largely maintaining a normal absolute CD3+ count in comparison to age-matched controls (Figure 3B). Clinical lymphocyte studies also indicated significant abnormalities in T cell naïve and memory subpopulations. The patient has persistently low proportion of CD4+ naive T cells and high proportion of CD4+ central memory and effector memory T cells. Additionally, the CD8+ T cell compartment shows an unusually high proportion of naive T cells, while the percentages of CD8+ central memory, effector memory, and TEMRA T cells are consistently low. This pattern underscores a persistent imbalance in the patient’s T cell maturation and memory formation processes. Memory B cells also showed a skewed distribution, with low levels of switched, unswitched memory and transitional B cells and high levels of naive B cells (Table S1).

**Figure 3.**
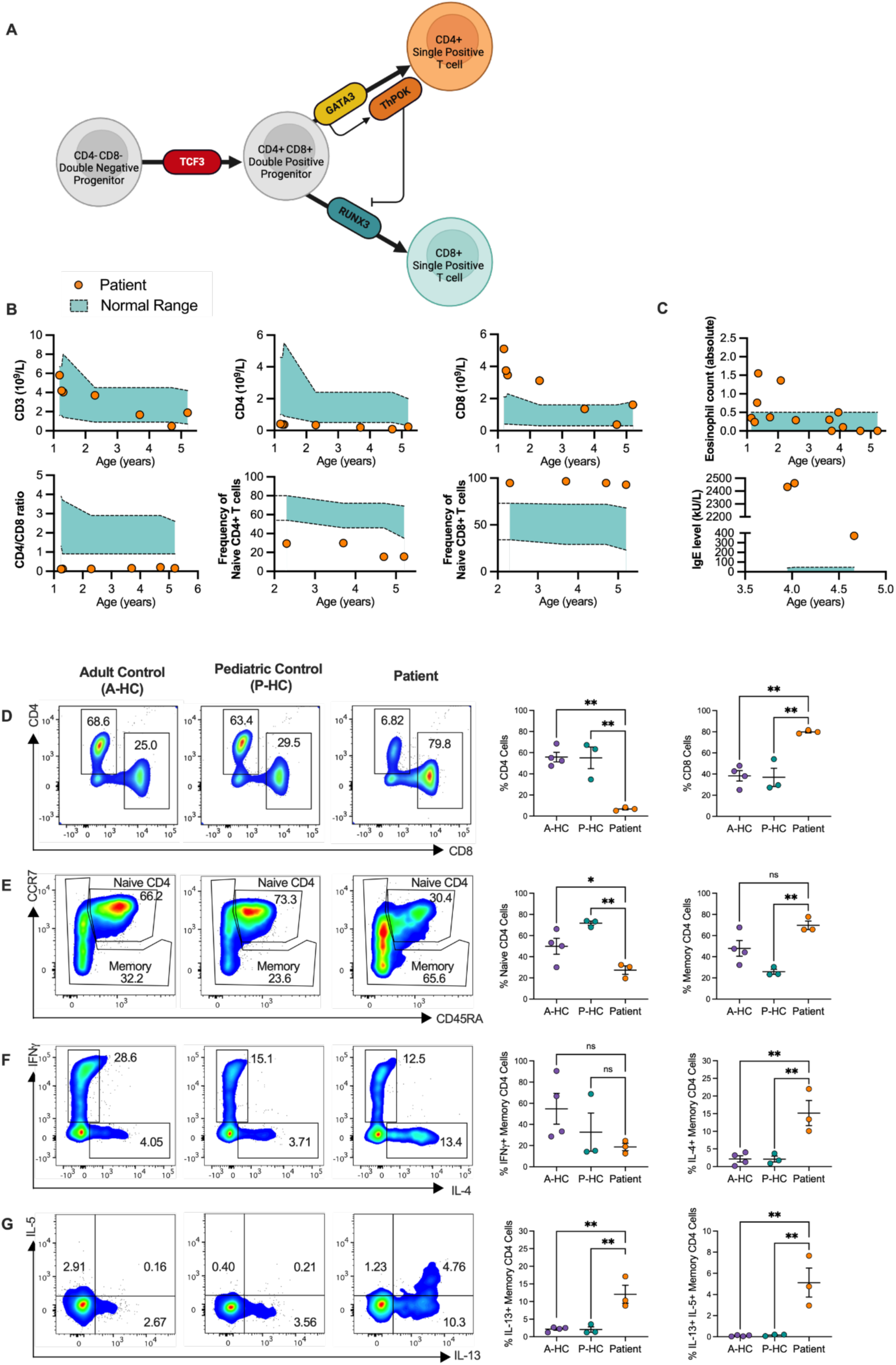
Patients CD4+ T cells exhibit impaired development and Th2 skewing post stimulation. **(A)** Schematic representation of ThPOK’s role in T cell lineage commitment. **(B)** Clinical flow cytometry of patient’s CD3+, CD4+, and CD8+ cells over time compared to pediatric healthy control (normal) range. **(C)** Absolute eosinophil count and serum IgE levels determined by the clinical laboratory over time. **(D)** Representative flow cytometry plots illustrating the proportions of CD4+ and CD8+ T cells in the patient compared to age-matched pediatric and adult healthy controls (HCs). HC, healthy control; A-HC, adult healthy control; P-HC, pediatric healthy control. Scatter dot plots summarize results from three independent blood draws for the patient compared to three age-matched pediatric (P-HC) and four adult HCs (A-HC). **(E)** Representative flow cytometry plots showing the proportions of memory and naive CD4+ T cells in the patient, identified by CCR7 and CD45RA markers, compared to age-matched pediatric and adult HCs. Scatter dot plots summarize results from three independent blood draws for the patient compared to three age-matched pediatric and four adult HCs. **(F-G)** Representative flow cytometry plots and analysis of Th2 cytokine production (IL-4, IL-5, and IL-13) in memory CD4+ T cells post-PMA/ionomycin stimulation. Scatter dot plots display comparative results. Data are mean ± SEM; statistical significance assessed by one-way ANOVA, with Dunnett’s method applied for multiple comparisons correction.

Along with the clinical observation of severe atopic manifestations in the patient, we also observed transient peripheral blood eosinophilia in the first 2 years of life and persistently elevated serum IgE levels (Figure 3C). This prompted an examination of the T helper 2 (Th2) cell in patient primary cells. We conducted an immunophenotyping study using flow cytometry to analyze the T cell compartment using blood drawn from the patient at three different points over two years. For comparative analysis, we included three age-matched pediatric healthy controls (HC) and four adult HCs. We first confirmed reduction in frequency of total CD4+ T cells and increase in CD8+ proportion in the CD3 compartment (Figure 3D). Within the CD4+ T cell population, there was an increase in the frequency of memory T cells alongside a substantial decrease in naive CD4+ T cells in the patient, in line with the clinically collected data (Figure 3E). Upon stimulation with PMA and ionomycin, a marked increase in Th2 cell activity was observed in the patient. This was characterized by elevated production of Th2 effector cytokines, including IL-4, IL-5, and IL-13 (Figure 3F-G). These findings suggest that the patient exhibits distinct immune dysregulation, particularly in the T cell compartment, with a pronounced shift towards memory phenotypes and Th2 effector function.

### Patient T cells exhibit impaired differentiation and activation

To understand the molecular underpinnings of the observed developmental T cell defect, we conducted single-cell RNA sequencing combined with antibody sequencing (scRNA-seq/Ab-seq) on the peripheral blood mononuclear cells (PBMCs) of the patient and two HCs, with and without T cell receptor (TCR) stimulation. Dimensionality reduction using UMAP on the unstimulated global transcriptomic signature did not place the patient’s T cells, NK cells, B cells or monocytes in a separate cluster from those of the HCs (Figure 4A). Quantitative assessment of T cell subset proportions, as measured using surface marker sequencing, did confirm clinical flow cytometry data: the patient exhibited low CD4+ T cells and high CD8+ T cells (Figure 4B). However, T cells specific clustering, visualized with UMAP, revealed that the patient’s T cells cluster separately from HC T cells. This difference in clustering was independent of CD4+, CD8+ or naïve and memory (using CD45RO) subset classification as determined by cell surface staining (Figure 4C). Patient T cells were largely naïve CD8+ but did not cluster with naïve T cells of HCs (Figure 4C).

**Figure 4.**
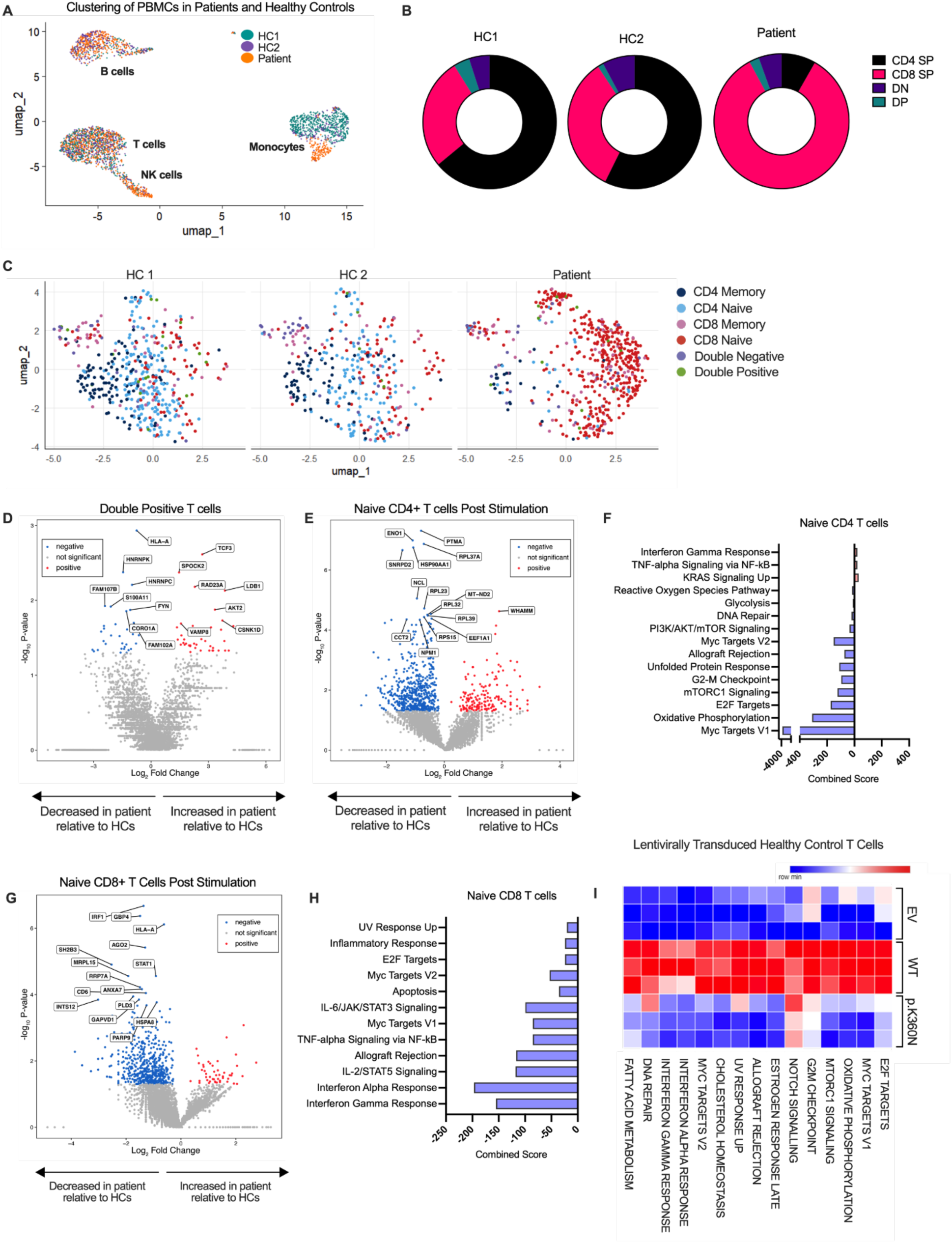
Patient T cells exhibit distinct clustering and gene expression profiles compared to healthy controls. **(A)** UMAP analysis of peripheral blood mononuclear cells from the patient and two age-matched healthy controls (HCs). HC, healthy controls; PBMC, peripheral blood mononuclear cells **(B)** scRNA-seq/Ab-seq analysis reflecting the proportion of CD4+ single positive, CD8+ single positive, double positive (DP) and double negative (DN) T cells in the patient and two HCs. **(C)** UMAP analysis of T cell clustering reveals distinct gene signatures between patient T cells and HCs. **(D)** Volcano plot comparing the differential expression of genes between DP cells of the patient in comparison to two age-matched HCs using scRNA-seq. **(E-H)** Transcriptome and Hallmark pathway analysis of post-TCR stimulated naïve CD4+ (in E and F) and CD8+ T cells (in G and H) in the patient compared to two age-matched HCs. **(I)** Pathway analysis of the differentially expressed genes assessed by RNA sequencing in HC T cells transduced with empty vector (EV), wild-type (WT), and p.K360N ThPOK.

Given the stark differences in transcriptomic signatures of patient T cells, we studied each of the key T cell subpopulations. Leveraging the BD® AbSeq Antibody-Oligonucleotides Conjugates, we were able to identify CD4+ and CD8+ expressing T cells in our scRNA-seq data. We examined double positive (DP) T cells, as they could reflect potential developmental defects in the final stages of T cell maturation in the thymus. We noted that the lineage commitment gene *TCF3* was upregulated in the patient’s DP T cells compared to HCs (Figure 4D). The accumulation of *TCF3* could suggest a defect in the downstream mediators of CD4+ T cell commitment, namely ThPOK and GATA3 (*9–11*). Subsequently, we investigated gene signatures in naïve CD4+ and CD8+ T cells. While we did not see differences between patient and HCs at baseline, we did see differences after TCR stimulation. Here, strong skewing was observed in both the naïve CD4+ and CD8+ compartment of patient T cells suggestive of impaired TCR activation compared to HCs (Figure 4E-H). A lack of T cell activation was particularly evident with patient cells failing to upregulate proliferative signaling pathways in CD4+ T cells (e.g., the MYC pathway) and effector CD8+ T cell pathways (e.g., interferon pathways) (Figure 4F and 4H). These results suggest a significant role for ThPOK in the activation of matured single-positive T cells. To establish this link experimentally, we stably transduced T cells with EV, WT, or p.K360N ThPOK containing lentiviral plasmids, and cultured them for 3 days. We observed that many pathways (e.g., MYC proliferation, and interferon effector) were upregulated in WT-transduced T cells, but these pathways failed to upregulate in p.K360N-transduced cells (Figure 3I). These results showed that the defects in T cell activation were attributable to the dominant-interfering effect of ThPOK^K360N^.

### ThPOK^K360N^ disrupts profibrotic gene suppression in pulmonary fibroblasts

In addition to the T cell defects, the patient carrying ThPOK^K360N^ had evidence of multi-organ fibrosis, and it has previously been shown that murine ThPOK binds to the regulatory regions of collagen genes in the skin to inhibit their expression (*4*). Using HEK293 cells, which express *COL2A1* but lack endogenous expression of ThPOK, we compared the effects of transduced ThPOK^K360N^ to ThPOK^WT^. Our results show that while ThPOK^WT^ suppresses *COL2A1* expression as expected, ThPOK^K360N^ does not, leading to significantly higher *COL2A1* levels similar to the control with an EV (Figure 5A). Additionally, cells expressing both ThPOK^WT^ and ThPOK^K360N^, mimicking a heterozygous state, also showed significantly increased *COL2A1* expression compared to ThPOK^WT^ alone (Figure 5A), indicating a dominant interfering effect of ThPOK^K360N^ on ThPOK^WT^ in repressing collagen expression. This led us to further validate our findings through stable lentiviral transduction, highlighting ThPOK’s critical role in regulating collagen gene expression. Since ThPOK is highly expressed in fibroblasts (www.proteinatlas.org/) and acts as a repressor of murine *Col1a1* and *Col1a2* (also highly expressed in fibroblasts) (*4*), we utilized primary human pulmonary fibroblasts as a model system. HC pulmonary fibroblasts were stably transduced with either EV, WT, or p.K360N ThPOK containing lentiviral plasmids (Figure 5B). Principal component analysis (PCA) of bulk RNA sequencing data showed clear and distinct clustering for each group. Principal Component 1 (PC1) and PC2 accounted for 52.6% and 32.9% of the total variance, respectively, and highlighted large differences in gene expression profiles influenced by ThPOK status (Figure 5C). Differential analysis between EV-transduced and WT ThPOK-transduced fibroblasts showed significant gene expression changes, many of which are not observed to the same level in variant-transduced cells (Figure 5D). Interestingly, overexpression of ThPOK^K360N^ led to altered gene expression profiles compared to ThPOK^WT^, with a number of genes that were upregulated in the WT being downregulated in the variant and vice versa (Figure 5D and 5E). This shift indicates that the missense change in the first zinc finger of ThPOK significantly changes its regulatory impact, potentially altering its DNA-binding affinity, interaction with other proteins, or responsiveness to cellular signals.

**Figure 5.**
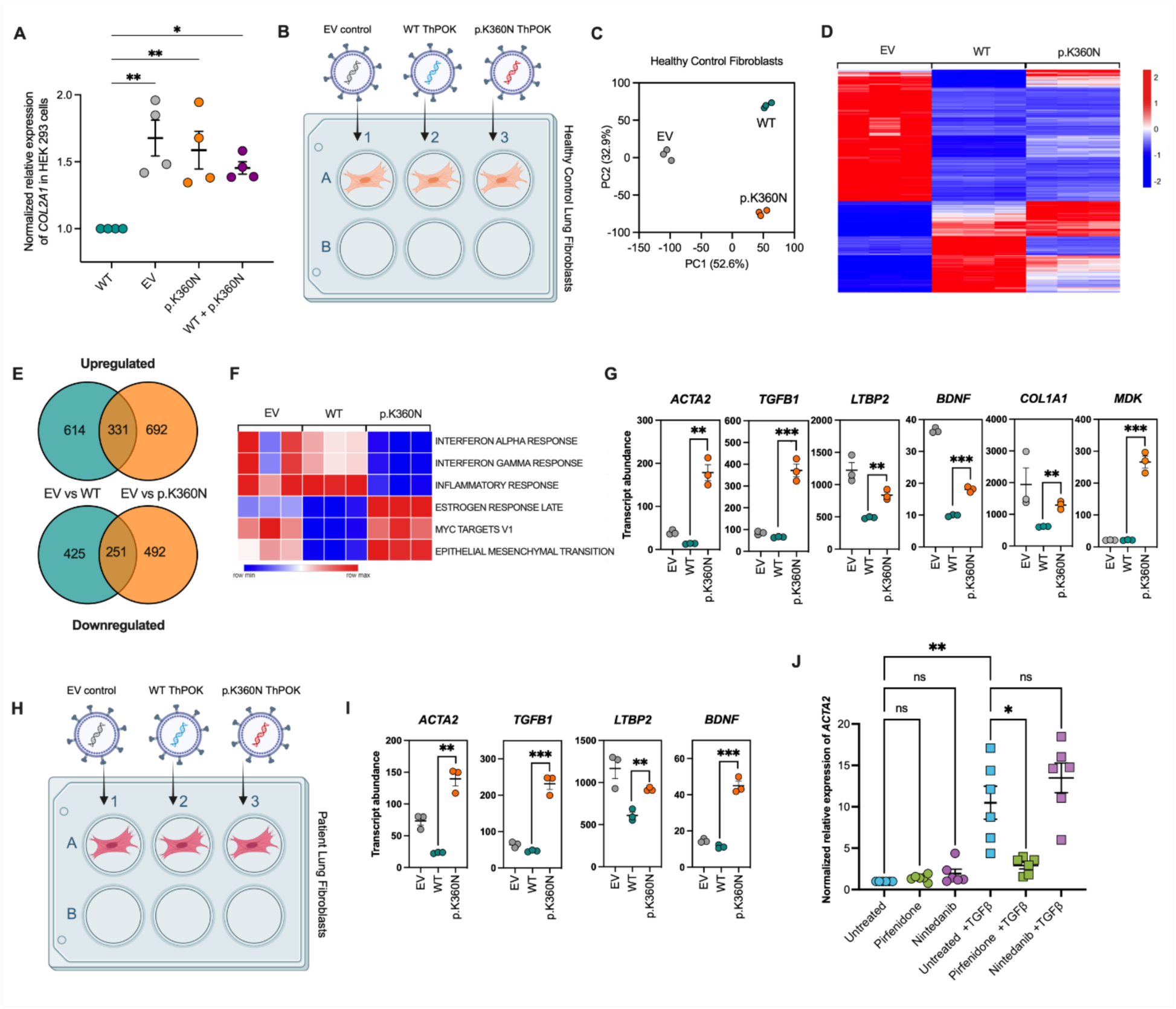
Expression of ThPOK^K360N^ impairs the suppression of profibrotic genes in primary pulmonary fibroblasts. **(A)** Expression of *COL2A1* measured by qPCR in HEK293 cells transfected with various constructs: empty vector (EV), wild-type (WT) ThPOK, p.K360N variant, or a combination of WT and the variant. Data are mean ± SEM; n=4 experiments; statistical significance assessed by one-way ANOVA, with Dunnett’s method applied for multiple comparisons correction. **(B)** Experimental schema depicting the transduction of healthy control primary human pulmonary fibroblasts. **(C)** PCA plot of healthy control primary human pulmonary fibroblasts transduced with WT ThPOK, p.K360N variant, or EV. **(D-E)** Heatmap and Venn diagram illustrating gene expression variations across all three groups in healthy control primary human pulmonary fibroblasts. Each group includes three biological replicates, with each replicate consisting of independently transduced, sorted, processed, and sequenced samples. **(F)** Sample Level Enrichment Analysis (SLEA) of transduced healthy control lung fibroblasts **(G)** Expression of profibrotic genes in transduced healthy control lung fibroblasts **(H)** Experimental schema showing patient-derived primary pulmonary fibroblasts, transduced with WT ThPOK, p.K360N variant, or EV. **(I)** Expression of profibrotic genes in transduced patient-derived primary lung fibroblasts **(J)** Expression of *ACTA2*, a known profibrotic marker, measured by qPCR to evaluate the effect of pirfenidone and nintedanib on profibrotic gene expression in TGF-β-treated patient-derived pulmonary fibroblasts. Data are mean ± SEM; n=6 experiments; statistical analysis was conducted using one-way ANOVA for repeated measures, with post-hoc comparisons performed using Fisher’s LSD test.

Pathway analysis of the transcriptomic data revealed downregulation of critical pathways including interferon alpha and gamma signaling, as well as the inflammatory response pathway in the variant-transduced cells compared to WT-transduced controls. Conversely, upregulation was observed in several pathways, such as estrogen response late, Myc targets, and epithelial mesenchymal transition (Figure 5F). The upregulation of the epithelial mesenchymal transition, a pathway integral to fibrosis and wound healing, was notable as it underlined the variant’s role in potentially exacerbating fibrotic conditions, highlighting a key area for further investigation. This is supported by the differential expression of several pro-fibrotic genes –*ACTA2* (*12–20*)*, TGFB1* (*18, 21, 22*)*, LTBP2* (*19, 23, 24*), and *BDNF* (*25*), and *COL1A1* (*20, 26*), and MDK (*27*)– that have been implicated in pulmonary fibrosis by multiple studies (Figure 5G). These genes have significantly increased transcript abundance in fibroblasts stably expressing ThPOK^K360N^ to compared to WT controls. These findings suggest that ThPOK^K360N^ represents a LOF in its ability to suppress fibrosis and profibrotic gene expression in fibroblasts, thereby highlighting the crucial role of ThPOK in maintaining cellular homeostasis against fibrotic processes.

### Reversal of fibrotic gene signature in patient pulmonary fibroblasts is achieved by over-expression of WT ThPOK or pirfenidone treatment

We next utilized patient-derived primary pulmonary fibroblasts to determine if stable overexpression of ThPOK^WT^ could reverse the observed fibrotic gene signature (Figure 5H). Our results revealed that the previously identified profibrotic genes in the transduced HC pulmonary fibroblasts exhibited similar differential regulation patterns in transduced patient-derived cells. Notably, there was a decrease in transcript abundance of the profibrotic genes in the patient-derived fibroblasts stably expressing the WT gene, thereby indicating a successful rescue of the patient-derived cells from the fibrotic phenotype (Figure 5I).

Building on our previous findings, we tested the efficacy of Food and Drug Administration (FDA)-approved pulmonary fibrosis drugs – pirfenidone (*28–30*) and nintedanib (*29–31*) – on patient-derived pulmonary fibroblasts in their steady state, and also under fibrosis-induced conditions. This approach was designed to better mimic the fibrotic microenvironment of the patient’s lungs, acknowledging that *in vivo*, cells are influenced by surrounding signals and are not isolated. For the induction of a fibrotic cell model, we employed transforming growth factor beta (TGF-β) stimulation, a method well-established in the literature for simulating a fibrosis response in human fibroblasts (*20, 30*). We used *ACTA2* expression via qPCR as our primary readout, due its recognition in the literature as a critical marker of myofibroblasts and activated fibroblasts, key players in fibrotic disease progression across various organs, including the lungs (*12–20*). Optimal *in vitro* treatment doses were determined by conducting dose-response experiments and assessing cytotoxicity in our model system using the lactate dehydrogenate (LDH) release cytotoxicity assay (Figure S7). We demonstrated that treatment with pirfenidone (1mM) over a 48-hour period reduced *ACTA2* expression in TGF-β-treated fibroblasts in comparison to untreated controls (Figure 5J). In contrast, a similar 48-hour treatment regimen with nintedanib (0.1μM) did not produce a comparable effect (Figure 5J). This finding highlights the efficacy of pirfenidone in modulating the fibrotic gene signature and may underscore the drugs’ distinct mechanisms of action and their varied impact on fibrotic pathways in patient-derived pulmonary fibroblasts.

## DISCUSSION

We describe the first reported human with a monogenic disorder caused by disruption of *ZBTB7B* which encodes the transcription factor ThPOK. This patient who was heterozygous for a variant in *ZBTB7B* presented with a complex syndromic phenotype including combined immunodeficiency, severe atopy, severe fibroinflammatory interstitial lung disease, sensorineural hearing loss, global developmental delay, and growth failure. Our investigations align with the established guidelines for genetic studies in single patients, as proposed by Jean-Laurent Casanova (*32*): the identified variant is not found in the unaffected parents or sibling; the variant is rare and absent in the gnomAD population database; *in silico* tools predict the variant to be damaging; and our functional investigations indicate that ThPOK^K360N^ exhibits multimorphic damaging effects.

The discovery of a monogenic defect in ThPOK in our patient has offered an exciting opportunity to explore the role of ThPOK in human immunology, providing a direct *in vivo* context that was previously inaccessible and primarily inferred from the HD mice and *in vitro* human studies. ThPOK is best known for its role as a ‘master regulator’ of CD4+ lineage commitment in the thymus, where it has been shown to be crucial for CD4+ versus CD8+ lineage decisions, particularly in MHC-II-restricted CD69+ CD4+ CD8^low^ intermediate thymocytes (*1–3, 8, 33–37*). This was first discovered through the HD mouse model characterized by the selective absence of mature CD4+ T cells in the periphery and increased numbers of CD8+ cells (*38*). Our observations of persistent CD4+ lymphopenia and CD8+ lymphocytosis in the patient’s T cell compartment recapitulate the phenotypic alterations seen in HD mice (*5, 36, 37*). HD mice are also noted to have markedly increased frequency of CD4+ CD8^low^ intermediate thymocytes (*38*). While this might suggest that class II-restricted thymocytes are arrested at this stage, ThPOK-deficient mice, including HD and knockout models, are reported to have MHC II-restricted CD8+ cells in both the thymus and spleen, suggesting that in the absence of functional ThPOK, MHC II-restricted thymocytes are reprogrammed to become CD8+ cells. This is supported by *in vitro* studies that show ThPOK-deficient CD4+ CD8^low^ thymocytes can differentiate into CD8+ cells (*1, 34*). This raises the question of whether all of the patient’s CD8+ cells are exclusively MHC I-restricted, or if the subset includes an MHC II-restricted component, an aspect that warrants further investigation.

Our study reveals significant differences in the clustering of T cells between the patient and HCs. While the patient’s T cells were predominantly naïve CD8+ cells, they did not align with the naïve CD8+ T cell clusters of HCs. This observation implies that the patient’s naïve CD8+ T cells possess unique characteristics or alterations that distinguish them from their healthy counterparts. Several factors could contribute to this unique clustering. Differences in TCR repertoires or altered signaling pathways in comparison to HC counterparts could all play a role in the distinct T cell profile of the patient. The patient’s immune environment might also influence T cell behavior and clustering. The altered clustering of naïve CD8+ T cells might also reflect changes in their activation potential, survival, or differentiation capacity. Further investigation is required to elucidate the specific factors driving the distinct clustering of the patient’s T cells.

In the context of CD4+ T cell biology, our findings complement the established literature by demonstrating that ThPOK not only orchestrates the development of CD4+ T cells in the thymus, but it also regulates the differentiation of several CD4+ T helper cell subsets and the activation and memory potential of both CD4+ and CD8+ T cells in the periphery (*36, 39, 40*). Notably, ThPOK plays a pivotal role in promoting Th2 cell differentiation while concurrently preventing the aberrant trans-differentiation of Th1/Th2 cells into cytotoxic T cell phenotypes (*36, 39*). Our data present an unexpected finding regarding Th2 cells. Contrary to existing reports that show the role of ThPOK in promoting Th2 cells (*36, 39*), the patient exhibited an increased Th2 response, with a significant enhancement in effector cytokine production (IL-4, IL-5, and IL-13) upon PMA/Ionomycin stimulation, suggesting an unanticipated role of ThPOK in modulating Th2 cell effector functions in humans. Our findings reveal an intriguing association between the observed increase in Th2 cell responsiveness and the patient’s severe atopic manifestations, which encompass early-onset and severe allergic conditions. While our data indicate a distinct Th2 cell hyperresponsiveness mediated by the variant, it remains unclear whether the enhanced Th2 responses are T cell intrinsic or influenced by other factors. It is possible that impaired regulatory T cell (Treg) differentiation or function, or altered basophil differentiation or function, may contribute to this phenotype. Additionally, assuming a T cell-intrinsic effect, the increased Th2 responses could stem from a loss of normal ThPOK function or the gain of new functions by the variant protein in the allergic inflammatory pathway.

ThPOK is also recognized to be important for CD4+ T cell activation, in particular preserving transcriptomic integrity and memory potential. In line with this, we observed a marked deficiency in the upregulation of genes essential for TCR-stimulated activation in naïve CD4+ T cells from the patient compared to HCs, highlighting a significant deficit in proliferative signaling pathways, such as the MYC pathway. The introduction of ThPOK^WT^ into T cells activated by TCR led to a notable upregulation of pathways critical for proliferation and effector functions, which were noticeably absent in cells transduced with the variant. Similarly, our research builds upon the known impact of ThPOK deficiency on clonal proliferation and effector molecule production in CD8+ T cells. ThPOK deficiency is shown to markedly impair the clonal proliferation and the production of CD8+ effector molecules, such as IL-2 and granzyme B, within long-lived CD8+ T memory (Tm) cells upon antigenic rechallenge (*41*). We identified a pronounced impairment in the activation of naïve CD8+ T cells in the patient, evidenced by a significant reduction in the upregulation of genes following TCR stimulation. This was particularly notable in the downregulation of effector pathways, such as those mediated by interferons, which are crucial for the functionality of CD8+ T cells. These findings suggest a direct link between ThPOK function and the activation of mature single-positive T cells. Our results thus provide an understanding of ThPOK’s role, not only in the differentiation and development of T cell subsets but also in their activation and functional response, thereby underscoring the complex regulatory mechanisms orchestrated by ThPOK in T cell immunobiology.

Beyond challenging the existing paradigm of ThPOK’s role in immunity, our study significantly advances the understanding of ThPOK’s function in human biology both mechanistically and clinically. Through transcriptomic analysis of transduced primary pulmonary fibroblasts, we uncovered substantial differences in pathway regulation between HC pulmonary fibroblasts expressing ThPOK^WT^ and those expressing the variant. Notably, we observed an upregulation in the epithelial-mesenchymal transition pathway, which is particularly relevant due to its association with fibrotic conditions. In addition, overexpression of ThPOK^K360N^ led to an exacerbated fibrotic gene signature, indicative of ThPOK’s involvement in regulating fibrotic processes beyond its involvement in the immune system. This was further evidenced by experiments with patient-derived pulmonary fibroblasts, where overexpression of ThPOK^WT^ reversed the fibrotic gene signature, pointing to ThPOK’s therapeutic potential in mitigating fibrotic diseases. Thus, our study suggests that genetic changes within ThPOK could have substantial consequences on the gene expression landscape in pulmonary fibroblasts, offering new insights into lung cell biology. Moreover, our exploration of FDA-approved pulmonary fibrosis drugs on patient-derived fibroblasts revealed that pirfenidone significantly reduced the expression of *ACTA2*, a marker of myofibroblasts and fibrosis, under both steady state and TGF-β-induced fibrotic conditions. The demonstration of pirfenidone’s ability to modulate the fibrotic gene signature in patient-derived fibroblasts is particularly promising as it suggests a possible therapeutic approach in managing the fibrotic aspects of diseases associated with ThPOK dysfunction. This finding not only emphasizes the value of targeted therapies in complex genetic diseases but also offers new insights into idiopathic pulmonary fibrosis and other fibrotic lung conditions. Additionally, echogenicity and functional abnormalities have been noted in the patient’s liver and kidneys, although these have not been definitively characterized as fibrosis. Given this newly characterized involvement of ThPOK in fibroblast regulation, it prompts speculation about the potential presence of fibrosis in other organs related to this variant. However, the exploration of such a hypothesis is constrained by the ethical and clinical limitations on obtaining biopsies without a clear medical indication from the patient.

In conclusion, here we describe a novel human disease associated with a multimorphic damaging variant in ThPOK, establishing its causative link to the patient’s clinical profile through complementary experimental approaches. This study not only broadens the scope of ThPOK’s known functions in immune regulation but also reveals its tissue-dependent dual regulatory nature, impacting both immune cells and fibroblasts. While our findings offer significant insights, they are derived from a single patient, the only identified case to date, highlighting a limitation and the need for further discovery of similar cases to expand the disease phenotype. Our research opens avenues for further research which includes assessing the impact of ThPOK variants across different tissues and their potential role in non-immune pathologies and a longitudinal study to track the patient’s disease progression. Moreover, further investigation into ThPOK’s interactions within the immune system, particularly its role in the skewed Th2 immune response, is essential. These future directions promise to deepen our understanding of ThPOK’s multifaceted role in human health and disease, paving the way for improved clinical management and therapeutic strategies for patients with ThPOK variant-associated conditions.

## MATERIALS AND METHODS

### Study design

The patient with a previously uncharacterized disease was identified after presenting to the immunology clinic. Trio whole-genome sequencing identified a *de novo* heterozygous variant in *ZBTB7B* that results in a missense change in the DNA-binding domain of ThPOK. We uncovered the altered DNA binding specificity and transcriptional activity of the variant by multiple approaches, including electrophoretic mobility shift assay (EMSA), high-throughput systematic evolution of ligands by exponential enrichment (HT-SELEX), luciferase assay, and chromatin immunoprecipitation followed by sequencing (ChIP-seq). We performed extensive phenotyping of the patient’s peripheral blood cells by single-cell RNA sequencing and conventional flow cytometry to reveal the abnormalities in T cell development, function and activation. To assess the mechanisms underlying the fibroinflammatory changes seen in the patient lung, we employed an *in vitro* system, utilizing lentiviral transduction to introduce both the wild-type (WT) and variant genes into primary pulmonary fibroblasts derived from healthy controls (HCs) and the patient. We also did treatment-testing using two Food and Drug Administration (FDA)-approved antifibrotic agents, one of which successfully reversed the fibrotic gene signature in patient primary pulmonary fibroblasts in an *in vitro* model of fibrosis.

### Study approval

The study participant and his parents/guardians and sibling provided written informed consent. Research study protocols were approved by the University of British Columbia Clinical Research Ethics Board (H15-00641).

### Genomic analysis

Genomic DNA from the patient and parents were sequenced with paired-end reads on the Ilumina platform by GeneDx. See supplementary methods for more details.

### Histopathology

A wedge biopsy was obtained from the edge of right upper lobe of the lung in a surgical open biopsy procedure. The immunohistochemistry stainings of the lung tissue were performed in BC Children’s Hospital Pathology Laboratory with clinically validated antibodies and protocols.

### Immunoblotting

Protein detection was carried out using standard immunoblotting techniques. Details are further elaborated in the supplemental methods. Briefly, HEK293 cells were cultured, transfected with relevant plasmids, lysed, and proteins were extracted. The proteins were separated by SDS-PAGE, transferred to membranes, and probed with anti-ThPOK (D9V5T) Rabbit mAb (Cell Signalling Technology, 1:1000), anti-Myc-Tag (9B11) Mouse mAb (Sigma Aldrich, 1:1000), and anti-β-Actin (8H10D10) Mouse mAb (Cell Signalling Technology, 1:20,000). Detection was performed using the Odyssey DLx Near-Infrared Fluorescence Imaging System.

### Luciferase reporter assay

A 1406 bp region of the promoter sequence of human *SOCS* (chr16:11,255,899-11,257,304 on GRCh38/hg38) was cloned into pGL4.20 [luc2/Puro] (Promega) firefly luciferase reporter plasmid. Detailed methods are provided in the supplemental section. Briefly, HEK293 cells were seeded in 24-well plates and incubated for 24 hours before transfection with EV, WT, or p.K360N variant plasmids, either individually (250 ng each) or in combination to model heterozygosity with varying ratios of WT to p.K360N (total 250 ng). Each well also received 250 ng of *SOCS1*-luciferase reporter and 10 ng PGL4.74 renilla luciferase control plasmid. Transfections were performed using Lipofectamine™ 3000. After 24 hours, cell lysates were prepared, and luciferase activity was measured using the Dual-Glo Luciferase Assay Kit (Promega) and a plate reader. In the analysis, firefly luciferase activity was normalized to renilla luciferase to account for transfection efficiency. This normalized activity was then divided by the value from the EV condition, providing a relative measure against the EV baseline.

### Electrophoretic mobility shift assay (EMSA)

Preparation of whole cell lysates was performed as previously described in the immunoblotting section above. A section of the *SOCS1* promoter, containing the WT ThPOK consensus DNA-binding sequence along with two variant sequences identified by HT-SELEX as preferentially bound by variant ThPOK, were used to design the double stranded oligonucleotide probes used in this assay (see Table S3 for sequences). Supershift assays were performed with 5 μg of whole cell protein lysate incubated on ice for 30 min with either anti-ThPOK (D9V5T) Rabbit mAb (Cell Signalling Technology, 1:1000) or Rabbit IgG Isotype Control (Invitrogen, Thermo Fisher Scientific) and then incubated at room temperature for 20 min with the labelled DNA probes and reagents provided in the Odyssey® EMSA Kit (LI-COR Biosciences) according to the manufacturer’s recommendations. Protein-oligonucleotide-antibody mixtures were then subjected to electrophoresis in 5% acrylamide/Bis-acrylamide 29:1 gel in 1x Tris-borate-EDTA (TBE) migration buffer (Thermo Scientific) for 90 min at 70 V at room temperature. Imaging was done using the Odyssey DLx Near-Infrared Fluorescence Imaging System (LI-COR Biosciences).

### High Throughput Systematic Evolution of Ligands by EXponential Enrichment (HT-SELEX)

WT and variant ThPOK (ZBTB7B_WT and ZBTB7B_K360N) were cloned into an eGFP expression vector pF3A-ResEnz-egfp. The TF samples were expressed by using a TNT SP6 High-Yield Wheat Germ Protein Expression System Kit (Promega). HT-SELEX was modified from our previous approach (*6*) to use Abcam Anti-GFP antibody ab290 immobilized to Protein G Mag Sepharose® Xtra Cytiva 28-9670-70) in the step where the protein–DNA complexes are separated from unbound DNA. The assay similarly used IVT-produced proteins, and the selection reactions were carried out in a buffer of 140 mM KCl, 5 mM NaCl, 1 mMK2HPO4, 2 mM MgSO4, 100 μM EGTA, 1 mM ZnSO4, and 20 mM HEPES-HCl (pH 7). After each of the three selection cycles, the ligands were amplified by PCR and output from all cycles was subjected to Illumina sequencing. Mung Bean Nuclease treatment also was used between each of the selection cycles to reduce the ssDNA background during the ligand selection. Data analysis was performed as previously described (*42*), where automatic detection of a sequence pattern defining local maxima was followed by semi manual generation of seeds that were then used to construct multinomial-1 or multinomial-2 position frequency matrices for the transcription factor target specificity.

### Chromatin Immunoprecipitation Sequencing (ChIP-seq)

To establish GFP-tagged ThPOK expressing HEK293 Flp-In-TRex cell lines, parental HEK293 Flp-In-TRex cells were transfected with separate expression vectors each carrying the WT or variant *ZBTB7B* open reading frames (FuGENE® HD Transfection Reagent, Promega) and after 48 hours transferred to Hygromycin selection media (0.2 ug/ul). Colonies of each line were pooled and used for further experiments. 24 hours prior to cross linking for chromatin immunoprecipitation, doxycycline (100 ng/ml) was added to cells and GFP expression was confirmed with fluorescent microscopy. Chromatin immunoprecipitation was performed as previously described (*43*). In brief, cells from 15 cm plates at 100% confluency were cross-linked for 10 min in 1% formaldehyde followed by 10 min of quenching with 2M glycine. After washing the cells with cold PBS, cells were collected and pelleted. Using a three-step lysis process, chromatin was released and then sonicated to produce DNA fragment length range of 200–300 bp using a Bioruptor sonicator (Diagenode). GFP-tagged proteins were immunoprecipitated with a polyclonal anti-GFP antibody (ab290, Abcam) and Dynabeads Protein G (Invitrogen). Crosslinks were reversed at 65°C overnight and bound DNA fragments were purified (QIAquick PCR Purification Kit, Qiagen). ChIP libraries were prepared using NEBNext Ultra II DNA kit. Sequencing was performed with 150n paired end at 2×10^7^ reads per sample. For each variant of *ZBTB7B*, two biological replicates and two input control samples were sequenced using NovaSeq 6000 Illumina sequencer.

In the data processing step, adapter sequences were trimmed from ChIP-seq reads with Cutadapt (v2.1) (https://doi.org/10.14806/ej.17.1.200) and mapped to the human genome (hg38) with Bowtie2 (v2.4.1) (*44*) using the –very-sensitive option. Reads with a map quality of less than 30 were discarded along with PCR duplicates and reads for which one or both of the paired ends could not be mapped. A single set of control reads was generated using the four-input control ChIP-seq replicates by merging them with SAMTools (v1.9) (*45*) and subsampling to one quarter of the total reads. MACS2 (v2.2.9.1) (*46*) was used to call peaks for each of WT and variant ThPOK ChIP-seq replicates using the merged input read set as the control. For each set of peaks, the top 2000 peaks with the highest enrichment scores were used for downstream analyses.

For each WT ThPOK ChIP replicate, we scanned the peak sequences to identify peaks that contained a match to the HT-SELEX WT ThPOK motif using FIMO (v5.5.0) (*47*) with default settings. Using BEDTools intersect (v2.30.0) (*48*), we overlapped the motif-containing ChIP peak summits for each replicate with a BED file containing the locations of human gene promoter regions (1000 bp upstream of the transcription start site to 500 bp downstream). To establish a set of high-confidence candidates for direct regulation by WT ThPOK, we took the intersection of genes identified from each replicate. We then performed the same analysis using the variant ThPOK ChIP replicates and the variant ThPOK HT-SELEX motif. We searched for overrepresented pathways in the set of high-confidence ThPOK regulated genes and variant ThPOK regulated genes using the PANTHER statistical overrepresentation test with PANTHER pathways (*49*). We checked each set in its entirety, each set excluding genes that overlapped with the other set, and the intersection of the two sets. We report all overrepresented PANTHER pathways with a false discovery rate P < 0.05.

### Isolation and culture of primary pulmonary fibroblasts

Fresh lung tissue was collected from the patient undergoing open lung biopsy for histopathological evaluation of interstitial lung disease. The tissue was immediately transported to the laboratory in ice-cold Dulbecco’s Modified Eagle Medium (Thermo Fisher Scientific) supplemented with 1% penicillin-streptomycin (Thermo Fisher Scientific). We followed an optimized published method for generating a fibroblast-enriched single-cell suspension combining mechanical and enzymatic dissociation. Concentrations for all the reagents used are reported in the original study (*50*). The processed cells were grown using Fibroblast Growth Medium 2 (PromoCell).

### Lentiviral transduction of primary pulmonary fibroblasts

To generate lentivirus vectors, WT, and p.K360N cDNA from the previously described expression plasmids were cloned into a GFP-tagged Lenti vector (cat #PS100071, OriGene) using EcoRI-HF and NotI-HF (New England BioLabs). The sequence-verified lentiviral plasmids were packaged using 3rd generation packaging plasmids and transfected into HEK293T cells using Lipofectamine™ 3000 Transfection Reagent (Thermo Fisher Scientific). Viral supernatant was harvested and filtered through a 0.45 μm PES filter (Thermo Scientific Nalgene) and concentrated using an Amicon Ultra-15 100 kDa centrifugal filter (Millipore). The concentrated lentivirus solution was then aliquoted and stored at −80°C until use. To establish patient-derived and healthy control-derived pulmonary fibroblasts that stably express WT and p.K360N, patient and healthy control cells were infected with EV, WT, or p.K360N lentiviral particles and 5ug/ml polybrene (Sigma-Aldrich) through spinfection at 1000 x g for 2 hours at 32°C, cultured, and expanded in Fibroblast Growth Medium 2 (PromoCell) for 3 days before undergoing sorting based on GFP expression using BD FACS Aria (BD Biosciences) cell sorter.

### RNA sequencing

To investigate the global transcriptome of the transduced patient and HC primary pulmonary fibroblasts stably expressing EV, WT, or p.K360N variant, 150,000 GFP+ cells were sorted based on GFP expression using BD FACS Aria (BD Biosciences) cell sorter directly into 500 ul of cold Buffer RLT Plus (Qiagen) supplemented with 2-Mercaptoethanol according to the Qiagen RNeasy Plus Mini Kit instructions. Following sorting, the volume of Buffer RLT Plus was adjusted so that there was exactly 350 ul of Buffer RLT Plus to 100 ul of sorted sample volume. RNA isolation was done according to the RNeasy Plus Mini Kit instructions (Qiagen). Sequencing was done on rRNA depleted RNA libraries using PE150 Illumina NovaSeq Sequencing, targeting 100M individual reads (50M read pairs/clusters) per sample. Three biological replicates were included for each condition, which were independently transduced, sorted, and sequenced.

Sequenced reads were aligned to a reference sequence using Spliced Transcripts Alignment to a Reference aligner. E1 Assembly and expression were estimated using Cufflinks E2 through bioinformatics apps on Illumina BaseSpace. Expression data was normalized to reads between samples using the edgeR package in R (R Foundation). Normalized counts were filtered to remove low counts using the filterByExpr function in edge. Principal component analysis (PCA) was done on log2 (normalized counts+0.25) in R using the PCA function. Differential expression analysis was accomplished using Limma. Differentially expressed genes were defined as those with adjusted p-value less than 0.05.

Pathway analysis was done by first performing Gene set enrichment analysis (GSEA) with 1000 permutations using the Molecular Signatures Database Hallmark module. Signal-to-noise ratio was used for gene ranking and the obtained P-values were further adjusted using the Benjamini-Hochberg method. Pathways with an adjusted P-value <0.05 were considered significant. Leading edge genes from significant pathways between WT and p.K360N-transduced cells were identified. Expression levels of these genes were then determined in each group.

Sample level enrichment analyses (SLEA) scores were computed for each significant pathway. Briefly, z-scores were computed for gene sets of interest for each sample. The mean expression levels of significant genes were compared to the expression of 1000 random gene sets of the same size. The difference between observed and expected mean expression was then calculated and represented on heatmaps generated using Morpheus (https://software.broadinstitute.org/morpheus).

### Single cell-RNA sequencing (scRNA-seq)

Using single cell RNA-Sequencing (scRNA-seq), whole transcriptome analysis (WTA) was conducted on peripheral blood mononuclear cells (PBMCs) from the patient and three age-matched healthy controls. The BD Rhapsody Single Cell platform was used, which included the Rhapsody Enhanced Cartridge Reagent Kit (BD), the BD Rhapsody Cartridge Kit (BD), the Rhapsody cDNA Kit (BD), the Rhapsody WTA Amplification Kit (BD), the Human Single-Cell Multiplexing Kit (BD), used according to manufacturer’s recommendations and protocols. Briefly, thawed PBMCs were rested overnight and then stimulated with ImmunoCult™ Human CD3/CD28 T Cell Activator (STEMCELL Technologies) or left untreated for 16 hours. Cells from each donor were then labelled with sample tags and a panel of BD® AbSeq Ab-oligos (BD), washed with stain buffer, and pooled together in cold sample buffer to obtain ∼60,000 cells in 620 ul for each of the pooled unstimulated and stimulated samples. Two nanowell cartridges were primed and subsequently loaded with the pooled samples. Libraries for whole transcriptome and AbSeq analysis were prepared by following the BD Rhapsody System (TCR/BCR Full Length, mRNA WTA, BD® AbSeq, and Sample Tag Library Preparation) Protocol. Quality control was performed using Agilent DNA High Sensitivity Kit (Agilent Technologies) and the Agilent 2100 Bioanalyzer (Agilent Technologies) on the intermediate and final sequencing libraries, including estimating the concentration of each sample, measuring the average fragment size of the libraries, and following sequencing recommendations. Libraries were diluted to 350-650 picomolar range per sample and sequenced using Illumina NovaSeq 150bp PE sequencing targeting 8,000M individual reads (4,000M read-pairs) with PhiX spike-in of 20%.

FASTQ files were processed using the BD Rhapsody Targeted Analysis Pipeline and Seven Bridges (www.sevenbridges.com) according to manufacturer’s recommendations. The R package Seurat was utilized for all downstream analysis. Scaling and clustering were performed on each pool of samples independently. Dimensionality reduction using PCA was done on the most variable genes, and UMAP was based on the first 20 PCs. Cell identities were annotated manually, or via cell surface antibody sequencing (Ab-seq), to first identify major cell types (T cells, B cells, NK cells and Monocytes) and then defining subtypes (CD4+ T cells, CD8+ T cells, Double Positive T cells, and their subsequent Naïve and Memory subgroups). For differential gene expression analyses, we utilized the Seurat implementation of negative binomial test, assuming an underlying negative binomial distribution in RNA-Seq data while leveraging the UMI counts to remove technical noise.

### Flow cytometry

To carry out immunophenotyping and intracellular cytokine detection, PBMCs from the patient and age-matched healthy controls were stimulated with eBioscience™ Cell Stimulation Cocktail (Invitrogen, Thermo Fisher Scientific) for 5 hours at 37°C. One hour after the start of stimulation, eBioscience™ Protein Transport Inhibitor Cocktail (Invitrogen, Thermo Fisher Scientific) was added to the stimulated cells. Cells were then stained with a cocktail of antibodies against surface markers for 20 minutes in room temperature and then fixed with Foxp3 Fixation/Permeabilization working solution from the eBioscience Foxp3 Transcription Factor Staining Buffer Set (Invitrogen, Thermo Fisher Scientific) for 20 minutes. The fixed cells were subsequently stained for 20 minutes with antibodies targeting intracellular cytokines in 1x Permeabilization Buffer (Invitrogen, Thermo Fisher Scientific). The samples were then washed and analyzed using the BD FACSymphony flow cytometer (BD Biosciences). Data were analyzed with FlowJo software (BD Biosciences). The antibody panels used for staining are listed in Table S4.

### Treatment testing and qPCR of primary fibroblasts

Primary patient pulmonary fibroblasts were used to evaluate the effects of various treatments. Initially, 5×10^5^ cells were plated in each well of a 6-well plate (Corning) containing Fibroblast Growth Medium 2 (PromoCell). The cells were incubated overnight under standard conditions. Subsequently, they were either left untreated or treated with recombinant human TGF-β1 (R & D Systems) at a concentration of 5 ng/μl for 24 hours to induce a fibrotic state, a well-established *in vitro* model of fibrosis as documented in existing literature. Following the TGF-β1 treatment, cells were subjected to treatment with Nintedanib (working concentration of 0.1 μM) or Pirfenidone (working concentration of 1 mM) for an additional 48 hours. Both drugs were purchased from MedChemExpress. For the groups designated to continue with TGF-β1 exposure, fresh medium supplemented with TGF-β1 was added concurrently with the drug treatments. This experimental setup allowed for the assessment of drug efficacy in both the presence and absence of an induced fibrotic environment. Post treatment, total RNA was extracted using a RNeasy Plus Mini Kit (Qiagen) and converted to cDNA using an iScript cDNA synthesis kit (BioRad Laboratories). Transcript abundance was measured using a Universal SYBR Green Super Mix (Bio-Rad) and a 7300 Real-Time PCR System (Applied Biosystems). Relative transcript abundance was quantified relative to Actin-β (*ACTB*) using the 2-ΔΔCT method. Pre-designed qPCR primers were purchased from Integrated DNA Technologies.

### Statistical Analysis

All data are presented as mean ± standard error of the mean (SEM). Statistical significance was evaluated using appropriate methods for each dataset which are detailed in the corresponding figure legends or the methods section, with the following annotations used to represent significance: p-val < 0.05 (*), p-val < 0.01 (**), p-val < 0.001 (***).

## Data Availability

All data produced in the present study are available upon reasonable request to the authors.

## Supplementary Materials

Materials and Methods

Figures S1 to S7

Tables S1 to S4

Data file S1

## Acknowledgments

We thank the patient and their family members for their participation in our study. We also acknowledge the BC Children’s Hospital BioBank for providing age-matched healthy control PBMC samples. Additionally, we thank Dr. Jonathan Bramson’s lab at McMaster University, Ontario, Canada, for providing the optimized lentivirus transduction protocol.

## Funding

Canadian Institutes of Health Research PJQ-173584 (SET)

Genome British Columbia SIP007 (SET)

Tier 1 Canada Research Chair in Pediatric Precision Health (SET)

Aubrey J. Tingle Professor of Pediatric Immunology (SET)

Health Professional-Investigator of the Michael Smith Foundation for Health Research (CMB)

Providence Healthcare Research Institute Early Career Investigator award (CMB)

Vanier Canada Graduate Scholarship (MV-S)

Four Year Doctoral Fellowship (MV-S, SS)

## Author contributions

Conceptualization: MV-S, MS, CMB, SET

Wet lab experiments: MV-S, MS, PY, SS, RR, AHWY, MB

Clinical data acquisition: CMB, SET, MV-S, PY, AFL, JHR, CLY, KJH, RB, MKD

Methodology: MV-S, MS, LDN, TRH, RB

Bioinformatic analyses: MS, MV-S, KUL, AJ, MA, PAR

Visualization: MV-S, MS, LG, KUL

Funding acquisition: CMB, SET

Supervision: CMB, SET

Writing – original draft: MV-S, MS

Writing – review & editing: All authors

## Data and materials availability

HT-SELEX and ChIP-sequencing data is available from ENA and SRA databases under accession: PRJEB75580.

