## Supplementary material for "A multimorphic variant in ThPOK causes a novel human disease characterized by T cell abnormalities, immunodysregulation, allergy, and fibrosis": ThPOK_manuscript_Supplementary Materials_2024.06.26.docx

**Methods and Materials**

**Genomic analysis**

Genomic DNA from the patient, mother and father were sequenced with paired-end reads on the Ilumina platform by GeneDx. Average mean sequencing coverage was reported to be at least 40x across the genome, with a minimum threshold of 30x for each sample. Bi-directional sequencing reads were assembled and aligned to the reference sequence based on NCBI Refseq transcripts and human genome build GRCh37/UCSC hg19. Using a custom-developed analysis tool (XomeAnalyzer), data were filtered and analyzed to identify sequence variants, repeat expansions, and most deletion and duplications greater than 1kb. Reported clinically significant variants were confirmed by an appropriate orthogonal method in the proband and the parents. A missense variant in *ZBTB7B* was reported as the only candidate with a potential relationship to the disease phenotype, which was only observed in the patient.

**Expression plasmid cloning**

WT Myc-DDK tagged *ZBTB7B* plasmid (cat #RC234388), corresponding to the human tagged open reading frame clone NM_001256455.2, was purchased from OriGene. The gene insert containing the point mutation corresponding to the patient’s variant (NM_001256455.2: c.1080A>C) was ordered from GenScript Biotech and inserted into the pCMV6-Entry plasmid (cat #PS100001, OriGene) using AsiSI and SacII restriction sites. The sequence integrity of the full-length plasmid was confirmed using long-read sequencing (Oxford Nanopore Technologies).

**Immunoblotting**

ThPOK expression was assessed by immunoblotting. Briefly, 1x10^6^ HEK293 cells were seeded in 6-well culture plates in 2 mL of Dulbecco modified Eagle medium (DMEM) with 10% FBS (Gibco, Life Technologies), 2mM L-glutamine (HyClone, Thermo Fisher Scientific), and 1mM sodium pyruvate (Gibco, Life Technologies) and incubated overnight at 37°C prior to transfection. Each well was transfected with 3 μg of relevant plasmids using Lipofectamine™ 3000 Transfection Reagent (Thermo Fisher Scientific) according to the manufacturer’s recommendations. Whole cell lysates were prepared 24 hrs post transfection by lysing cells in RIPA Lysis and Extraction Buffer (Thermo Fisher Scientific) supplemented with the Halt protease and phosphatase inhibitor cocktail (Thermo Fisher Scientific). The protein concentrations were measured using Pierce™ Coomassie Plus Assay Reagent (Thermo Fisher Scientific). Laemmli Sample Buffer (Bio-Rad Laboratories) supplemented with β-mercaptoethanol was added to the cell lysates and boiled at 37°C for 5 minutes. Lysates were separated by 10% SDS-PAGE and transferred onto polyvinylidene difluoride membranes (Bio-Rad Laboratories). Membranes were blocked using 5% BSA in Tris-buffered saline supplemented with Tween-20, incubated with anti-ThPOK (D9V5T) Rabbit mAb (Cell Signalling Technology, 1:1000), anti-Myc-Tag (9B11) Mouse mAb (Sigma Aldrich, 1:1000), and anti-β-Actin (8H10D10) Mouse mAb (Cell Signalling Technology, 1:20,000) primary antibodies in blocking buffer overnight at 4˚C, and lastly incubated with anti-rabbit (IgG DyLight 800, Rockland Immunochemicals) and anti-mouse (IgG IRDye 680RD, LI-COR) secondary antibodies at a concentration of 1:20,000 for 1hr at room temperature in blocking buffer. The membranes were imaged using the Odyssey DLx Near-Infrared Fluorescence Imaging System (LI-COR Biosciences).

**Luciferase reporter assay**

A 1406 bp region of the promoter sequence of human *SOCS* (chr16:11,255,899-11,257,304 on GRCh38/hg38) was cloned into pGL4.20 [luc2/Puro] (Promega) firefly luciferase reporter plasmid using KpnI and HindIII restriction sites. The sequence integrity of the full-length plasmid was confirmed by long-read sequencing (Oxford Nanopore Technologies). Briefly, 1.5x10^5^ HEK293 cells were seeded in 24-well culture plates in 0.5 mL of DMEM with 10% FBS, 2mM L-glutamine, and 1mM sodium pyruvate and incubated at 37°C for 24 hours prior to transfection. We analyzed the effects of the EV, WT, and p.K360N variant plasmids, both individually and in combination, on the activity of the *SOCS1* promoter. To test the effect of each plasmid in isolation, cells were transfected with 250 ng of the EV, WT, or the p.K360N variant plasmid. To model the heterozygous state, co-transfections were conducted where cells received a combination of WT and p.K360N plasmids in increasing ratios of WT to p.K360N (0.5:1, 1:1, 2:1, 4:1, and 8:1) totaling to 250 ng. Additionally, all cells were transfected with 250 ng of *SOCS1*-luciferase reporter plasmid and 10 ng of PGL4.74 renilla luciferase control plasmid (Promega). The transfection was carried out using Lipofectamine™ 3000 (Thermo Fisher Scientific) according to the manufacturer’s protocol. After 24 hours, cell lysates were prepared using 1x Glo Lysis Buffer (Promega) and transferred to white flat-bottom 96-well plates in technical triplicates. Dual-Glo Luciferase Assay Kit (Promega) was used according to manufacturer’s recommendations and luciferase activity was measured using the Infinite M200 plate reader (Tecan) by integrating luminescence over 10 seconds per well. In the analysis step, the firefly luciferase activity was normalized against renilla luciferase which controlled for variation in transfection efficiency. The normalized firefly luciferase activity was further divided by the normalized value from the EV condition, providing a relative measurement against the EV baseline.

**Structural predictions**

Due to the absence of a crystallized structure for ThPOK (encoded by *ZBTB7B*), we utilized the known crystal structure of its closely related homolog, ZBTB7A. This homolog shares significant similarity with ThPOK (*ZBTB7B*), especially in the four-finger DNA-binding domain and the C2H2 zinc finger domain, making it a suitable proxy for our analysis. Using ChimeraX-1.6, we visualized a schematic of the interaction of the lysine residue in ZBTB7A (analogous to the lysine at position 360 in the WT ThPOK) or the variant residue (p.K360N) with DNA.

**Treatment testing and qPCR of primary fibroblasts**

Primary patient pulmonary fibroblasts were used to evaluate the effects of various treatments. Initially, 5x10^5^ cells were plated in each well of a 6-well plate (Corning) containing Fibroblast Growth Medium 2 (PromoCell). The cells were incubated overnight under standard conditions. Subsequently, they were either left untreated or treated with recombinant human TGF-β1 (R & D Systems) at a concentration of 5 ng/μl for 24 hours to induce a fibrotic state, a well-established in vitro model of fibrosis as documented in existing literature. Following the TGF-β1 treatment period, cells were subjected to different drug treatments for an additional 48 hours. These treatments which were purchased from MedChemExpress ([www.medchemexpress.com](http://www.medchemexpress.com)) which included Nintedanib (0.01μM, 0.1μM, 1μM, and 10μM) and Pirfenidone (0.25mM, 0.5mM, 1mM, and 2mM). For the groups designated to continue with TGF-β1 exposure, fresh medium supplemented with TGF-β1 was added concurrently with the drug treatments. Post treatment, total RNA was extracted from treated and untreated cells using a RNeasy Plus Mini Kit (Qiagen) and converted to cDNA using an iScript cDNA synthesis kit (BioRad Laboratories). Transcript abundance was measured using a Universal SYBR Green Super Mix (Bio-Rad) and a 7300 Real-Time PCR System (Applied Biosystems). Relative transcript abundance was quantified relative to Actin-β (*ACTB*) using the 2-ΔΔCT method.

**Lactate dehydrogenase release assay for assessing cytotoxicity**

Cytotoxicity was assessed using the LDH-Glo™ Cytotoxicity Assay (Promega Corporation) according to the manufacturer’s recommendations. This assay quantitatively measures lactate dehydrogenase (LDH) release from damaged cells into the culture medium. Supernatants were obtained from cells that undergone treatment testing (outline above). At the end of the treatment period, 5 µL of supernatant from each well was diluted with 495 µL of LDH storage buffer, achieving a 100x dilution. Subsequently, 50 µL of this diluted sample was transferred to a new 96-well plate. To each well, 50 µL of reconstituted LDH detection reagent was added. The plates were incubated at room temperature for 30 minutes, protected from light. Luminescence was measured using the Infinite M200 plate reader (Tecan) by integrating luminescence over 1000 milliseconds (ms) per well. Percent cytotoxicity was determined by comparing the LDH release from experimental wells to a maximum LDH release control. The control consisted of cells lysed with 40 µL of 10% Triton X-100 in a total medium volume of 2000 µL. The LDH release from each experimental well was divided by this maximum LDH release value to calculate the percent cytotoxicity. Additionally, a standard dilution curve of LDH, ranging from 32 mU/mL to 0 mU/mL, was employed to confirm that the measured values fell within the assay’s reliable detection range.

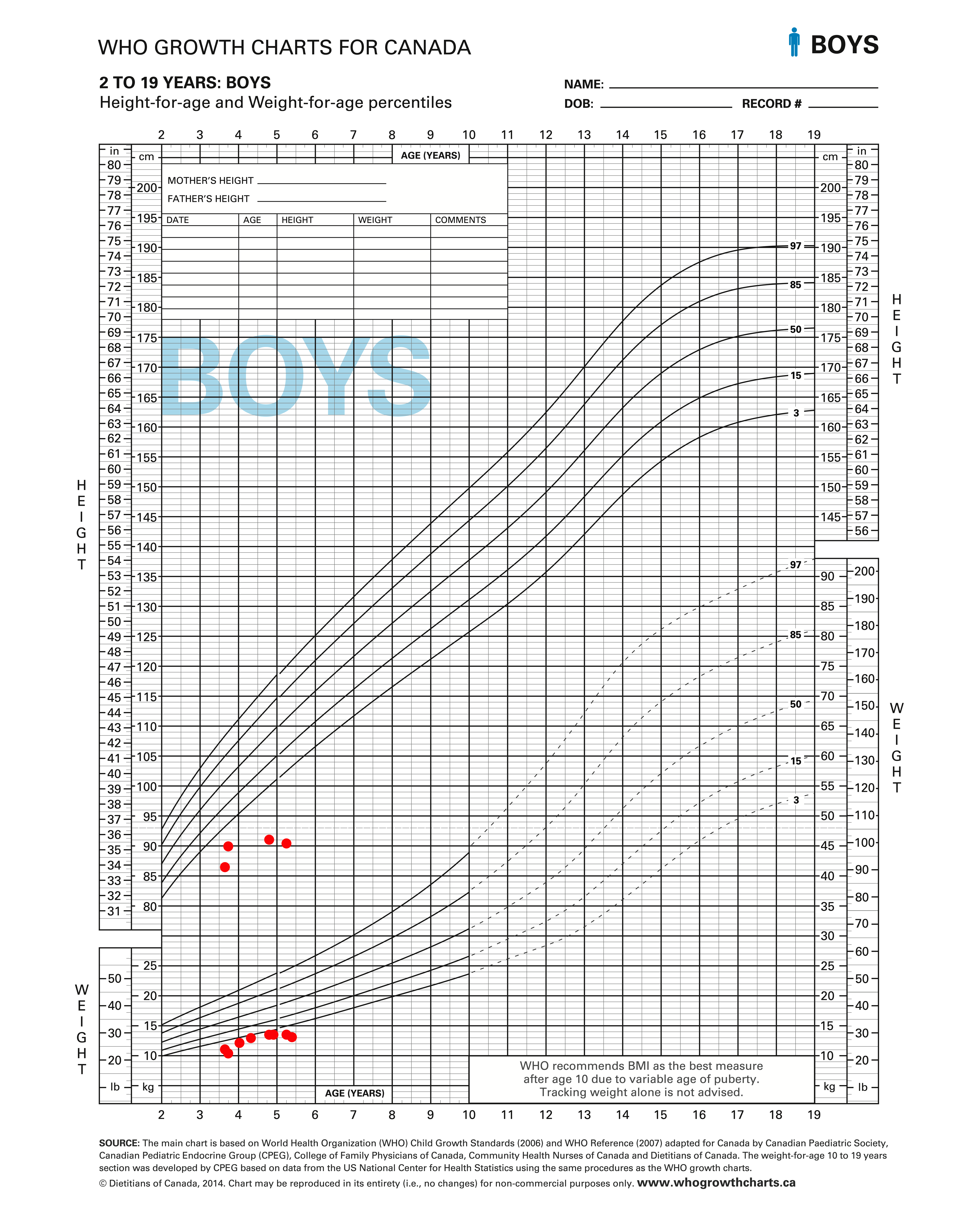

**Figure S1. Growth chart for height and weight of the patient.** This chart adapted for Canadian boys between the ages of 2 to 19 from the World Health Organization (WHO) displays the height and weight measurements of the patient over a two-year period. Both parameters are consistently below the 3rd percentile, indicating significantly lower growth compared to typical developmental milestones for children of the same age group.

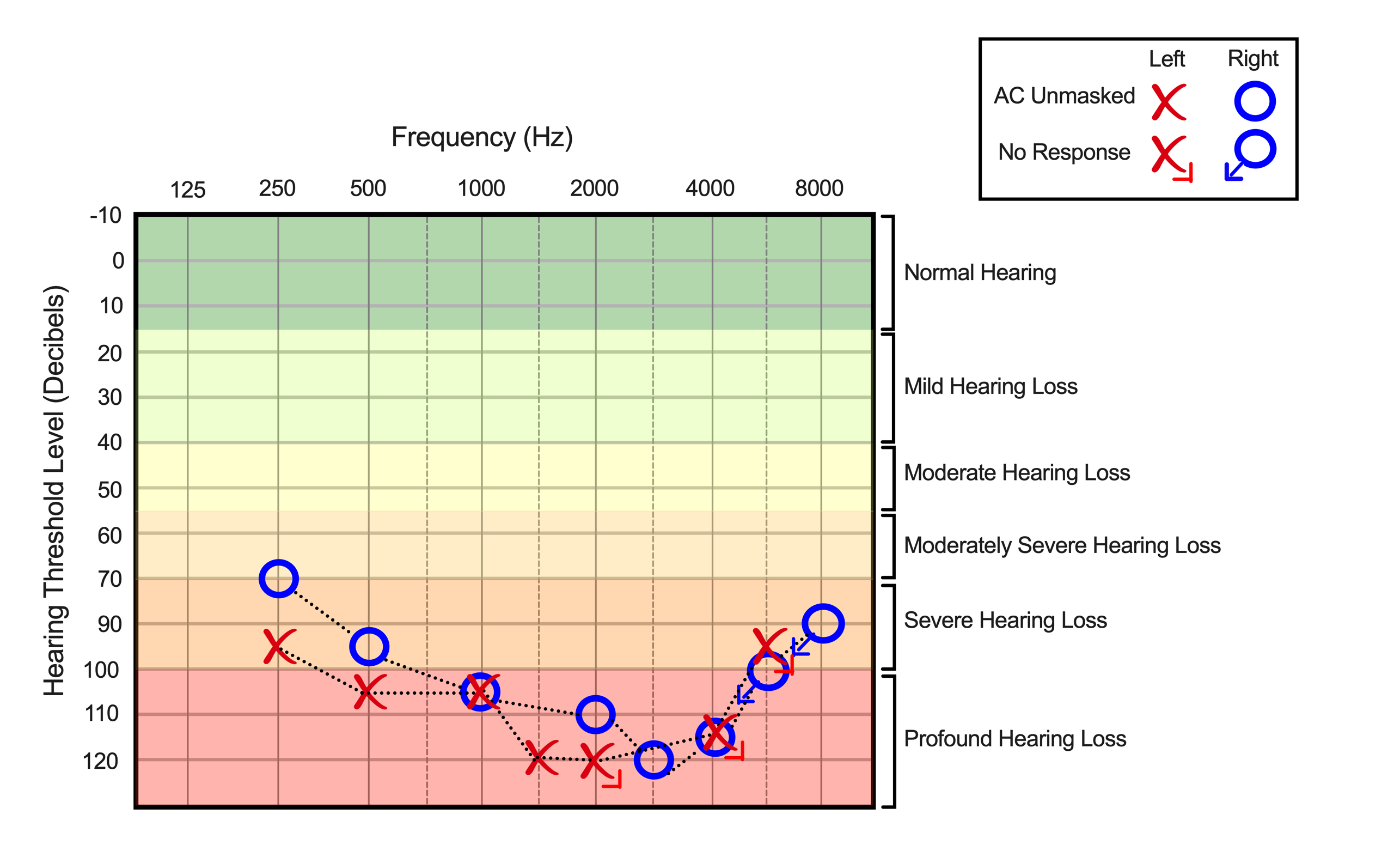

**Figure S2. Auditory Brainstem Response (ABR) testing results**. The testing was conducted using warble tone stimuli through conditioned play audiometry with insert earphones and bone-conductor, both demonstrating excellent reliability. This figure summarizes the results from the ABR testing indicating severe to profound sensorineural hearing loss in both ears, with more pronounced loss in the left ear. For the left ear, severe to profound sensorineural hearing loss was recorded (95 - 120 dB HL) from 250 - 1500 Hz, with no responses at higher frequencies (2000 - 8000 Hz). For the right ear, severe to profound sensorineural hearing loss was recorded (70 - 120 dB HL) from 250 - 4000 Hz, with no responses at higher frequencies (6000 - 8000 Hz).

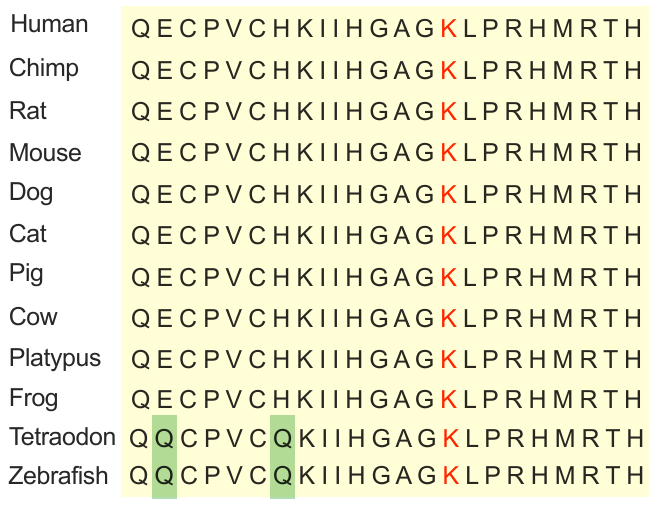

**Figure S3. Conservation of the lysine residue across species.** In the amino acid sequence of the first C2H2 zinc finger of ThPOK, Lysine (K) is consistently present in all 12 species examined, highlighting it as a highly conserved amino acid across these diverse organisms.

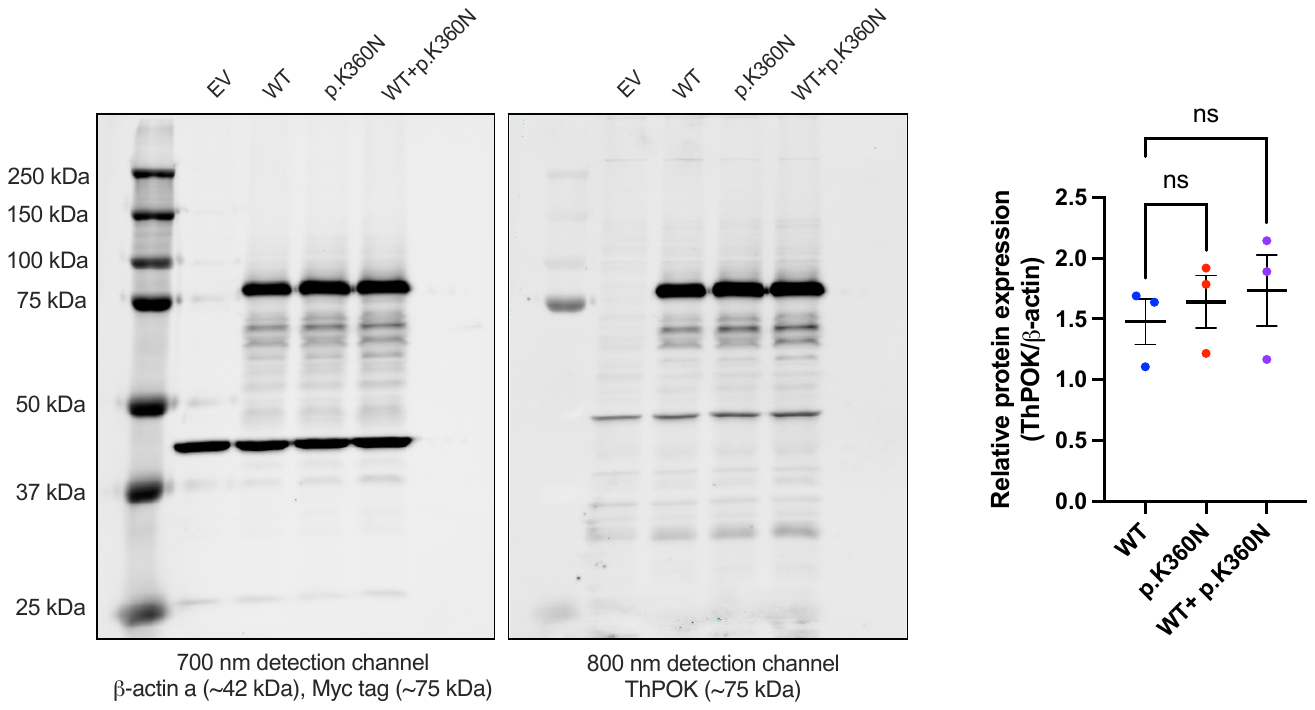

**Figure S4. Full-length immunoblots and quantification of ThPOK expression.** On the left, full-length immunoblots of lysates obtained from HEK293 cells transfected with plasmids expressing wild-type ThPOK (WT), p.K360N ThPOK (p.K360N), or an empty vector (EV) control. The immunoblots were probed with antibodies against ThPOK, Myc-tag, and β-actin. Equal loading was demonstrated with the anti-β-actin antibody. Blots are representative of three independently repeated experiments. On the right, quantification of band intensities was done using Empiria Studio® Software (Licor). Data presented are mean ± SEM from three independent experiments. One-way ANOVA was used for statistical analysis with Dunnett's multiple comparisons test to account for multiple comparisons; ns, not significant.

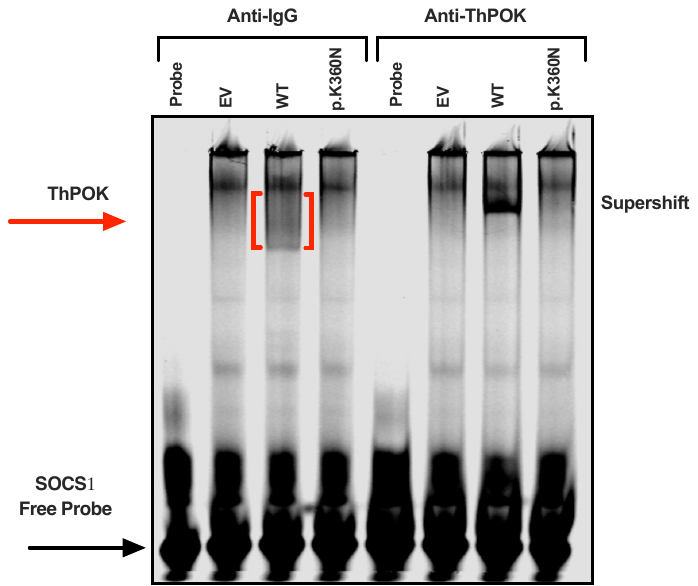

**Figure S5. Electrophoretic mobility shift assay (EMSA) to assess binding of the wild-type and variant ThPOK proteins to known wild-type ThPOK consensus sequence.** HEK293 cells were transfected with plasmids expressing wild-type ThPOK (WT), p.K360N ThPOK (p.K360N), or an empty vector (EV) control. Whole cell lysates were analyzed for binding to wild-type (WT) ThPOK consensus sequence (CGACCACC) contained in the promoter of *SOCS1*. Anti-IgG antibody (on the left) was used as a control to confirm the specificity of the observed supershift in the ThPOK-DNA complex (on the right). Image is representative of three independently repeated experiments. Refer to Table S3 for the sequence of the DNA probe used in this assay.

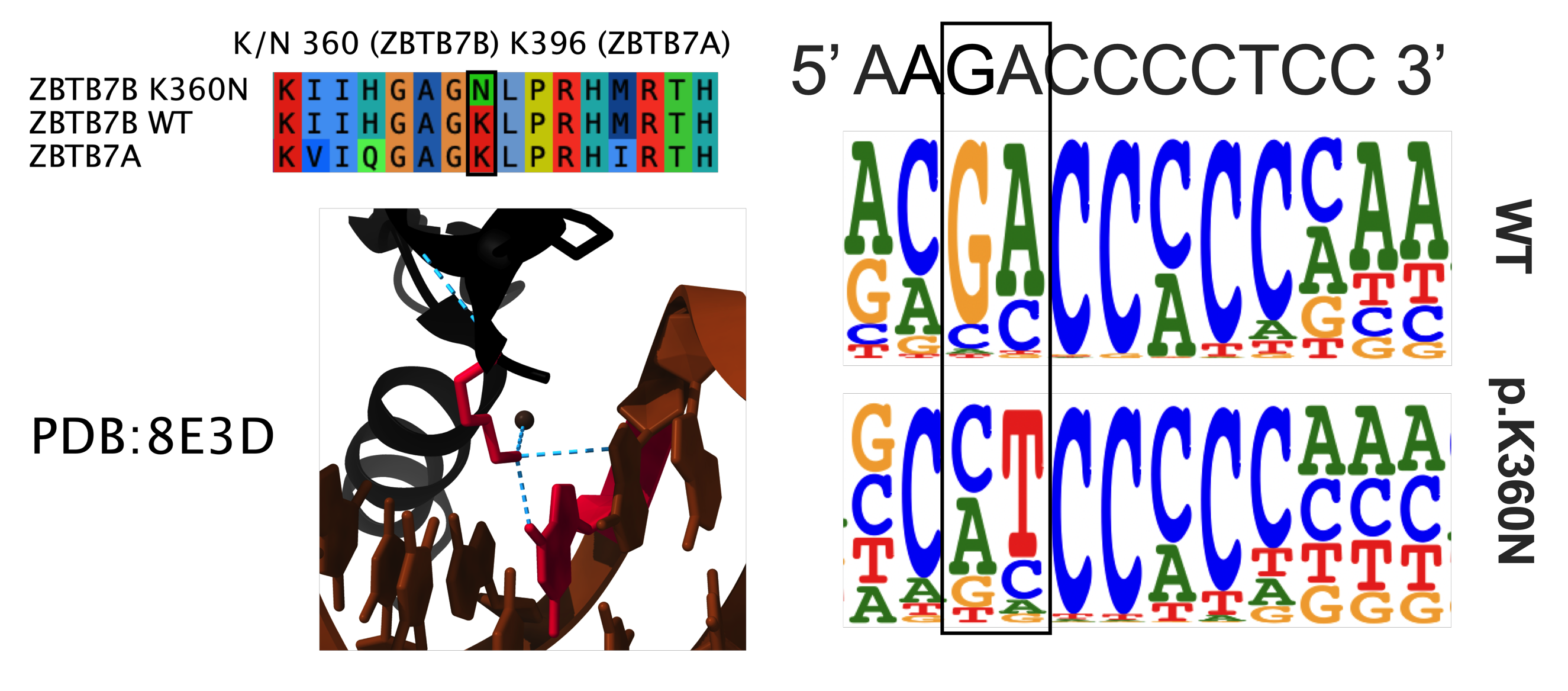

**Figure S6. Structural modeling analysis of ThPOK interaction with DNA.** This figure uses the crystal structure of the homologous protein ZBTB7A (PDB: 8E3D) to predict the impact of an amino acid substitution at position 360 in ThPOK. ZBTB7A shares sequence homology with ThPOK in the DNA-binding zinc finger domain (*1*). The structural analysis indicates that in ZBTB7A, a lysine residue—corresponding to position 360 in ThPOK—forms essential hydrogen bonds with the GA dinucleotide. This suggests that the amino acid change at position 360 in ThPOK likely disrupts these interactions, potentially altering its DNA binding specificity.

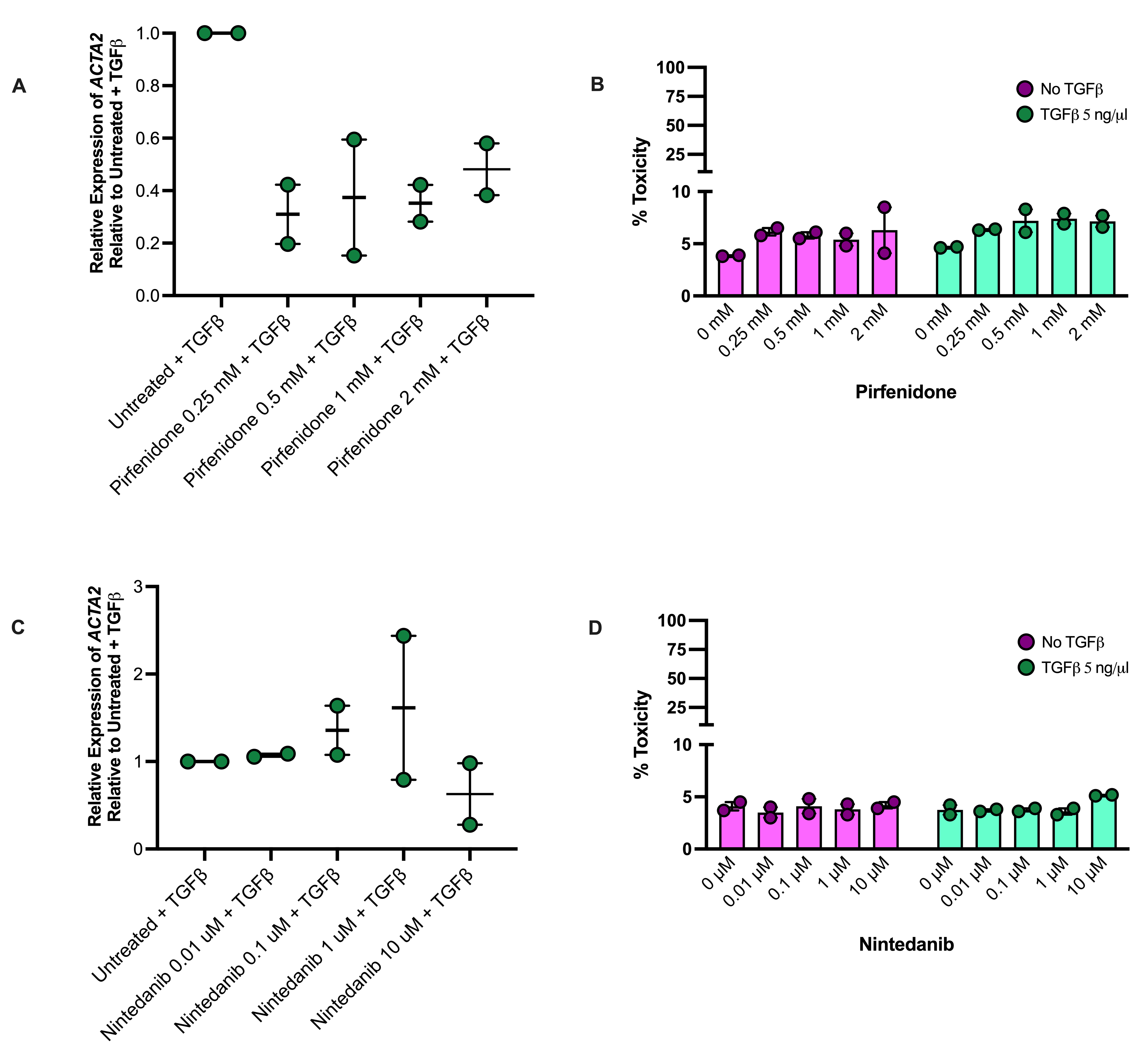

**Figure S7. Evaluating the efficacy and toxicity of pirfenidone and nintedanib in TGF-β-treated patient primary pulmonary fibroblasts.** (A and C) Dose-response graphs illustrating the expression of *ACTA2*, a profibrotic marker, in response to four different doses of pirfenidone or nintedanib. Expression levels were measured using qPCR to evaluate the effect of pirfenidone or nintedanib on profibrotic gene expression in patient-derived pulmonary fibroblasts treated with recombinant human TGF-β1 (5 ng/µL for 24 hours) to induce fibrosis. Following TGF-β1 induction, cells were exposed to varying concentrations of pirfenidone or nintedanib for an additional 48 hours. Data represent mean ± SEM from two independent experiments. (B and D) Cytotoxicity associated with different doses of pirfenidone or nintedanib, assessed by lactate dehydrogenase (LDH) release assay using supernatants from A and C.

**Table S1. Clinical flow cytometry studies of the patient from 1 to 5 years old.**

|  | **Patient** | **Reference** | **Patient** | **Reference** | **Patient** | **Reference** |
| --- | --- | --- | --- | --- | --- | --- |
| **Complete blood count differential** | | | | | | |
| White blood cells (x10^9^/L) | 17.5 (H) | 5.0-15.0 | ----- | ----- | 17.8 (H) | 5.3-16.0 |
| Neutrophils (x10^9^/L) | 13.5 (H) | 1.50-8.50 | ----- | ----- | 6.45 | 1.50-8.50 |
| Lymphocytes (x10^9^/L) | 3.1 | 2.00-8.00 | ----- | ----- | 10.15 | 4.00-10.50 |
| Monocytes (x10^9^/L) | 0.7 | 0.00-0.90 | ----- | ----- | 0.36 | 0.00-0.90 |
| Eosinophils (x10^9^/L) | 0.0 | 0.00-0.50 | ----- | ----- | 0.76 (H) | 0.00-0.50 |
| Basophils (x10^9^/L) | 0.1 | 0.00-0.20 | ----- | ----- | 0.06 | 0.00-0.20 |
| **Lymphocyte studies** | | | | | | |
| CD3+cells (x10^9^/L) | 1.89 | 0.70-4.20 | 3.71 | 0.90-4.50 | 4.19 | 1.60-6.70 |
| CD3+ CD4+ cells (x10^9^/L) | 0.23 (L) | 0.30-2.00 | 0.34 (L) | 0.50-2.40 | 0.37 (L) | 1.00-4.60 |
| CD3+ CD8+ cells (x10^9^/L) | 1.61 | 0.30-1.80 | 3.11 (H) | 0.30-1.60 | 3.74 (H) | 0.40-2.10 |
| CD3- CD56+ cells (x10^9^/L) | 0.33 | NA | 0.64 | NA | 0.14 | NA |
| CD19+cells (x10^9^/L) | 0.65 | 0.20-1.60 | 2.81 (H) | 0.20-2.10 | 3.37 (H) | 0.60-2.70 |
| **T cell naïve, memory, and effector** | | | | | | |
| CD4+ Naïve T cells (% CD4+ T cells) | 15.5 (L) | 35.0-69.0 | 29.4 (L) | 54.0-80.0 | ----- | ----- |
| CD4+ Central Memory T cells (% CD4+ T cells) | 61.6 (H) | 9.0-25.0 | 53.3 (H) | 10.0-26.0 | ----- | ----- |
| CD4+ Effector Memory T cells (% CD4+ T cells) | 22.1 | 10.0-30.0 | 16.7 (H) | 3.0-16.0 | ----- | ----- |
| CD4+ TEMRA T cells (% CD4+ T cells) | 0.8 (L) | 4.0-22.0 | 0.6 (L) | 3.0-12.0 | ----- | ----- |
| CD57+ cells (% CD4+ T cells) | 1.0 | NA | 0.8 | NA | ----- | ----- |
| PD-1+ cells (% CD4+ T cells) | 49.2 | NA | 34.0 | NA | ----- | ----- |
| CD8+ Naïve T cells (% CD8+ T cells) | 92.9 (H) | 23.0-68.0 | 94.8 (H) | 34.0-73.0 | ----- | ----- |
| CD8+ Central Memory T cells (% CD8+ T cells) | 3.7 (L) | 4.0-11.0 | 1.8 (L) | 3.0-15.0 | ----- | ----- |
| CD8+ Effector Memory T cells (% CD8+ T cells) | 1.4 (L) | 14.0-59.0 | 1.3 (L) | 9.0-47.0 | ----- | ----- |
| CD8+ TEMRA T cells (% CD8+ T cells) | 2.0 (L) | 6.0-30.0 | 2.2 (L) | 7.0-25.0 | ----- | ----- |
| CD57+ cells (% CD8+ T cells) | 1.3 | NA | 0.2 | NA | ----- | ----- |
| PD-1+ cells (% CD8+ T cells) | 2.6 | NA | 1.7 | NA | ----- | ----- |
| **B cell memory** | | | | | | |
| Switched memory B cells (% B cells) | 0.1 (L) | 4.0-16.0 | ----- | ----- | ----- | ----- |
| Unswitched memory B cells (% B cells) | 0.9 (L) | 5.0-16.0 | ----- | ----- | ----- | ----- |
| Naïve B cells (% B cells) | 95.2 (H) | 65.0-86.0 | ----- | ----- | ----- | ----- |
| Transitional B cells (% B cells) | 2.2 (L) | 9.0-24.0 | ----- | ----- | ----- | ----- |
| CD21^low^ B cells (% B cells) | 1.9 (L) | 3.0-10.0 | ----- | ----- | ----- | ----- |

**Table S2. Variant description and *in silico* predictors of pathogenicity.**

| **Gene** | *ZBTB7B* (Zinc finger and BTB domain containing 7B) |
| --- | --- |
| **Chromosome** | Chromosome 1 |
| **Genomic position (GRCh37)** | Chr1(GRCh37):g.154988216A>C |
| **cDNA position (MANE Select, RefSeq)** | NM_001256455.2(ZBTB7B):c.1080A>C |
| **Protein variant (NP_001186796.1)** | NP_001243384.1:p.Lys360Asn (K360N) |
| **Exon affected** | Exon 2 |
| **Zygosity** | Heterozygous |
| **Inheritance** | *De novo* |
| **CADD (v1.6)** | 23.7 |
| **PolyPhen2** | Probably damaging (score: 0.999) |
| **SIFT (v6.2.0)** | Deleterious |
| **gnomAD** | Variant not found |

**Table S3. List and sequence of IRDye labelled WT and variant probes used in EMSA.**

| SOCS1 promoter probe | 5’caaattacagccc**CGACCACC**gaccgcccaccatc 3’ | sense – IRDye700 |
| --- | --- | --- |
|  | 5’ gatggtgggcggtcggtggtcggggctgtaatttg 3’ | anti-sense |
| Variant Probe 1 | 5’ caaattacagccc**CATCCCCC**gaccgcccaccatc 3’ | sense – IRDye700 |
|  | 5’ gatggtgggcggtcgggggatggggctgtaatttg 3’ | anti-sense |
| Variant Probe 2 | 5’ caaattacagccc**CCTCCACC**gaccgcccaccatc 3’ | sense – IRDye700 |
|  | 5’ gatggtgggcggtcggtggagggggctgtaatttg 3’ | anti-sense |

The bolded letters within the sequences represent the core DNA-binding consensus sequence. The dinucleotides highlighted in green and red denote regions corresponding to wild-type (WT) and variant dinucleotide changes, respectively, as identified by high-throughput systematic evolution of ligands by exponential enrichment (HT-SELEX).

**Table S4. List of antibodies used in the T cell phenotyping research flow cytometry panel.**

| **Marker** | **Fluorophore** | **Dilution** | **Catalog #** | **Company** |
| --- | --- | --- | --- | --- |
| Viability | eF780 | 1:100 | 65-0865-14 | Invitrogen |
| CD3 | BUV805 | 1:100 | 612896 | BD Horizon |
| CD4 | BUV563 | 1:100 | 612923 | BD Horizon |
| CD8 | BV570 | 1:100 | 301037 | Biolegend |
| CD45RA | PerCP-Cy5.5 | 1:100 | 563429 | BD Pharmingen |
| CCR7 | APC-R700 | 1:100 | 565868 | BD Horizon |
| IFN-gamma | BV605 | 1:50 | 562974 | BD Horizon |
| TNF-alpha | BUV395 | 1:50 | 563996 | BD Horizon |
| IL-4 | PE | 1:50 | 554485 | BD Pharmingen |
| IL-5 | BV421 | 1:50 | 504311 | Biolegend |
| IL-13 | PE-Cy7 | 1:50 | 501914 | Biolegend |
| IL-17A | AlexF488 | 1:10 | 560488 | BD Pharmingen |
| IL-21 | AlexF647 | 1:10 | 560493 | BD Pharmingen |
